# Durability of mRNA-1273 against COVID-19 in the time of Delta: Interim results from an observational cohort study

**DOI:** 10.1101/2021.12.13.21267620

**Authors:** Ana Florea, Lina S. Sy, Yi Luo, Lei Qian, Katia J. Bruxvoort, Bradley K. Ackerson, Gina S. Lee, Jennifer H. Ku, Julia E. Tubert, Yun Tian, Carla A. Talarico, Hung Fu Tseng

## Abstract

**Background:** We conducted a prospective cohort study at Kaiser Permanente Southern California to study the vaccine effectiveness (VE) of mRNA-1273 over time and during the emergence of the Delta variant.

**Methods:** The cohort for this planned interim analysis consisted of individuals aged ≥18 years receiving 2 doses of mRNA-1273 through June 2021, matched 1:1 to randomly selected unvaccinated individuals by age, sex, and race/ethnicity, with follow-up through September 2021. Outcomes were SARS-CoV-2 infection, and COVID-19 hospitalization and hospital death. Cox proportional hazards models were used to estimate adjusted hazard ratios (aHR) with 95% confidence intervals (CIs) comparing outcomes in the vaccinated and unvaccinated groups. Adjusted VE (%) was calculated as (1-aHR)x100. HRs and VEs were also estimated for SARS-CoV-2 infection by age, sex, race/ethnicity, and during the Delta period (June-September 2021). VE against SARS-CoV-2 infection and COVID-19 hospitalization was estimated at 0-<2, 2-<4, 4-<6, and 6-<8 months post-vaccination.

**Results:** 927,004 recipients of 2 doses of mRNA-1273 were matched to 927,004 unvaccinated individuals. VE (95% CI) was 82.8% (82.2-83.3%) against SARS-CoV-2 infection, 96.1% (95.5-96.6%) against COVID-19 hospitalization, and 97.2% (94.8-98.4%) against COVID-19 hospital death. VE against SARS-CoV-2 infection was similar by age, sex, and race/ethnicity, and was 86.5% (84.8-88.0%) during the Delta period. VE against SARS-CoV-2 infection decreased from 88.0% at 0-<2 months to 75.5% at 6-<8 months.

**Conclusions:** These interim results provide continued evidence for protection of 2 doses of mRNA-1273 against SARS-CoV-2 infection over 8 months post-vaccination and during the Delta period, and against COVID-19 hospitalization and hospital death.

**Summary:** This prospective cohort study provides evidence for continued protection of 2 doses of mRNA-1273 against SARS-CoV-2 infection over 8 months post-vaccination and during the Delta period. VE against COVID-19 hospitalization remained robust and stable over the same period.

## Introduction

Since coronavirus disease 2019 (COVID-19) vaccine rollout, multiple observational studies have demonstrated that mRNA-1273 (Moderna Inc, Cambridge, USA) is highly effective against symptomatic COVID-19, with reports of vaccine effectiveness (VE) as high as 100% [1–4]. mRNA-1273 has also been shown to have high VE against COVID-19 hospitalization and death [1, 3].

However, the emergence of several SARS-CoV-2 variants led to concerns of potentially lower VE of COVID-19 vaccines against variants [5, 6]. The higher transmissibility of the Delta variant led to a surge in infections, hospitalizations, and deaths in the United States (US) since becoming the dominant circulating strain in July 2021 [7, 8]. Even though a majority of cases were in unvaccinated individuals, breakthrough cases among vaccinated individuals have also been reported [9–12]. We previously evaluated the VE of mRNA-1273 against SARS-CoV-2 variants, and reported an overall VE of 86.7% against Delta infection through July 2021 [13]. Another test-negative observational study in Ontario, Canada, from December 2020 to August 2021, reported a 94% VE against symptomatic infection with Delta after two doses of mRNA-1273 with similar VE against hospitalization and death [14]. Furthermore, several studies conducted in the US post Delta emergence reported high VE of mRNA-1273 against COVID-19 hospitalization ranging from 91.6% to 95.0% [15–17].

As the pandemic continues, the durability of protection of COVID-19 vaccines, for which data are limited [18], and the potential need for boosters have become critical public health questions. We previously reported that waning of VE was most pronounced for the Delta variant, with VE against Delta infection declining from 94.1% at 14-60 days after vaccination to 80.0% at 151-180 days after vaccination [13]. A test-negative study conducted in the United Kingdom compared VE of mRNA-1273 against Delta symptomatic disease over 14 weeks, showing a decline in VE from 95.2% at 1 week after vaccination to 90.3% at 14 weeks after vaccination [19].

Few published studies are available on the durability of protection of mRNA-1273 several months after vaccination during the Delta period among the general US population. In this study, we evaluated the durability (with up to 8 months of follow-up) and effectiveness of 2 doses of mRNA-1273 in preventing SARS-CoV-2 infection, stratified by age, sex, and race/ethnicity, and during the Delta period in the US. We also evaluated VE and durability against COVID-19 hospitalization.

## Methods

### Study setting

Kaiser Permanente Southern California (KPSC) is an integrated health care system providing care to a diverse 4.6 million members[20]. KPSC’s electronic health record (EHR) is comprehensive, capturing details across several areas such as inpatient, outpatient, emergency, and virtual care; claims also capture any care received outside of the KPSC system. The KPSC Institutional Review Board reviewed and approved the study.

### Study objectives

This is the second planned interim analysis (IA) of a 5-year cohort study at KPSC to evaluate the VE of mRNA-1273 in preventing SARS-CoV-2 infection and severe COVID-19 disease. Detailed methods and results of the first IA with follow-up through June 30, 2021 were reported elsewhere [1]. For this analysis, the primary objectives were to evaluate the VE of 2 doses of mRNA-1273 in preventing SARS-CoV-2 infection and severe COVID-19 disease. Secondary objectives were to evaluate the VE of 2 doses of mRNA-1273 in preventing SARS-CoV-2 infection by age, sex, and race/ethnicity groups. We also evaluated the waning of VE over time to assess the durability of mRNA-1273 against SARS-CoV-2 infection and COVID-19 hospitalization. To specifically assess VE against Delta in the absence of waning, we also evaluated VE against Delta infection in individuals newly vaccinated in June 2021 and followed through September 30, 2021. The study protocol is presented in the Supplementary Materials.

### Study population

For the study cohort, eligible individuals were ≥18 years old, and were KPSC members for at least 1 year prior to the index date (allowing a 31-day membership gap) and for ≥14 days after the index date. Individuals who received 2 doses of mRNA-1273 ≥24 days apart (4-day grace period allowed prior to the recommended 28-day interval) from December 18, 2020 through June 30, 2021 made up the vaccinated group. The index date was defined as the date of receipt of the second dose of mRNA-1273 for vaccinated individuals; their matched unvaccinated counterparts were assigned the same index date. Individuals who received a COVID-19 vaccine other than mRNA-1273 on or before the index date, or who had no healthcare utilization or no vaccination from the 2 years prior to the index date through the index date were excluded. Individuals who died, had a COVID-19 outcome, or received any COVID-19 vaccine □14 days after the index date were also excluded.

The unvaccinated group was composed of those who did not receive any COVID-19 vaccine as of the index date. Unvaccinated comparators were randomly selected and matched 1:1 to the vaccinated individuals by age group (18–44 years, 45–64 years, 65–74 years, and ≥75 years), sex, and race/ethnicity (Non-Hispanic White, Non-Hispanic Black, Hispanic, Non-Hispanic Asian, and Other/Unknown). Index dates were also balanced through matching, since matched vaccinated and unvaccinated individuals shared the same index date.

### Exposure and outcomes

Information on mRNA-1273 exposure was ascertained from KPSC’s EHR. The EHR included vaccines administered within KPSC as well as COVID-19 vaccines administered outside of the KPSC health system. These were imported daily into the KPSC EHR from the California Immunization Registry (CAIR), CalVax (Cal Poly Pomona mass vaccination site), Care Everywhere (Epic EHR feature which allows health care systems to exchange medical information), claims (e.g., pharmacies), and member self-report with documentation. All providers of COVID-19 vaccines were required by law to provide COVID-19 vaccine administration data daily to CAIR [21].

The first primary outcome was SARS-CoV-2 infection defined as a positive molecular test or a COVID-19 diagnosis code for both symptomatic and asymptomatic infections. Individuals with COVID-19 who had a history of a COVID-19 diagnosis code or a SARS-CoV-2 positive molecular test in the 90 days prior were not considered incident cases. The second primary outcome was severe COVID-19 disease, which included COVID-19 hospitalization (hospitalization with a SARS-CoV-2 positive test or a COVID-19 diagnosis code, or a hospitalization ≤7 days after a SARS-CoV-2 positive test) and COVID-19 hospital death (death occurring during COVID-19 hospitalization). COVID-19 hospitalization was confirmed 1) by at least one documented SpO_2_ <90% during hospital stay, or 2) if SpO_2_ ≥90%, by manual chart review done by a physician investigator [BKA] and trained chart abstractors to verify the presence of severe COVID-19 symptoms.

Individuals were followed for COVID-19 outcomes ≥14 days after the index date through September 30, 2021 (end of follow-up) or until occurrence of a censoring event (termination of KPSC membership allowing for a 31-day gap, death, or receipt of a COVID-19 vaccine), resulting in up to 8 months of follow-up post-vaccination for this analysis. Unvaccinated individuals stopped contributing unvaccinated person-time if they received a first dose of mRNA-1273 during follow-up and started contributing vaccinated person-time when they received an eligible second dose of mRNA-1273.

Starting from June 1, 2021, Delta (B.1.617.2, AY.*) was the predominant strain among positive SARS-CoV-2 specimens successfully sequenced at KPSC [1]. To evaluate VE of mRNA-1273 specifically against SARS-CoV-2 infection during the Delta period, a subset of individuals was included in an additional Delta period analysis if they had an index date during June 1-30, 2021 and were followed up to September 30, 2021.

### Other variables

Baseline characteristics were extracted from the EHR. Variables assessed at index date included age, sex, race/ethnicity, socioeconomic status (Medicaid coverage, neighborhood median household income), medical center area, pregnancy status, and KPSC physician/employee status. Variables assessed in the two years prior to index date included smoking and body mass index (BMI). Variables assessed in the year prior to index date included Charlson comorbidity score, autoimmune conditions, health care utilization (virtual, outpatient, emergency department [ED], inpatient encounters), preventive care (other vaccinations, screenings, well-visits), chronic diseases (kidney disease, heart disease, lung disease, liver disease, diabetes), and frailty index [22]. Other variables included history of SARS-CoV-2 infection and molecular test performed from March 1, 2020 to index date (irrespective of result), and immunocompromised status.

### Statistical analyses

Baseline characteristics of the vaccinated and unvaccinated groups were described. Continuous variables were compared using two-sample t-test or Wilcoxon rank-sum test, as appropriate, and categorical variables were compared using χ^2^ test or Fisher’s exact test, as appropriate. Potential confounders were identified based on the literature. To assess the balance of covariates across groups, absolute standardized differences (ASD) were used. Covariates with ASD>0.1, as well as age, sex, race/ethnicity, month of index date were included in the multivariable models. The missing indicator method was used for covariates with missing data [23].

The number of incident events divided by person-years was used to calculate the incidence rates (IR) per 1,000 person-years of SARS-CoV-2 infection, and COVID-19 hospitalization and hospital death for both vaccinated and unvaccinated groups. Kaplan-Meier curves were used to estimate the cumulative incidence of SARS-CoV-2 infection, and COVID-19 hospitalization and hospital death for both groups; the differences between the curves were tested by log-rank test.

Cox proportional hazards regression models were used to estimate unadjusted and adjusted hazard ratios (HR) with 95% confidence intervals (CIs) comparing SARS-CoV-2 infection, and COVID-19 hospitalization and hospital death in the vaccinated and unvaccinated groups overall. VE (%) was calculated as (1-HR) x 100. HRs and VEs were also estimated comparing SARS-CoV-2 infection in vaccinated and unvaccinated individuals by age, sex, race/ethnicity, and during the Delta period.

To evaluate durability of protection, VE against SARS-CoV-2 infection and COVID-19 hospitalization was estimated every two months post-vaccination (0-<2, 2-<4, 4-<6, and 6-<8 months) using time-varying Cox regression models. VEs by months post-vaccination against SARS-CoV-2 infection and COVID-19 hospitalization were also estimated and stratified by age groups (18-64 years old and ≥65 years old).

All analyses were conducted using SAS software version 9.4, Cary, USA.

## Results

### Baseline characteristics

The study cohort consisted of 927,004 recipients of 2 doses of mRNA-1273 vaccine (hereafter, ‘vaccinated’) and 927,004 matched unvaccinated individuals (**Figure 1**). Overall, the median age was 52 years (interquartile range [IQR] 36-65) with 74.3% aged 18-64 years; there were more females than males (54.5%); and 38.6% were Hispanic, 33.6% were non-Hispanic White, 12.8% were Asian, and 7.5% were Black (**Table 1**). The vaccinated and unvaccinated groups had similar characteristics (ASD≤0.1) of BMI, smoking, Charlson comorbidities index, frailty index, chronic diseases, immunocompromised status, autoimmune conditions, pregnancy status, history of SARS-CoV-2 infection and molecular test, ED visits, hospitalizations, Medicaid enrollment, and median household income. Compared to the unvaccinated individuals, those vaccinated had more outpatient and virtual visits, and more preventive care visits in the year prior to the index date. A higher proportion of vaccinated individuals were KPSC physicians/employees. There were some differences in the distribution of vaccinated and unvaccinated groups across the medical centers. The greatest proportion of index dates occurred in April 2021 (34.5%) followed by May 2021 (21.2%), coinciding with California’s vaccination phase when mRNA-1273 became available to all individuals ≥18 years old [24]. Vaccinated individuals received their second dose after a median 28 days (IQR 28-29) post-first dose, and 0.1% received a concomitant vaccine with mRNA-1273.

**Table 1.**
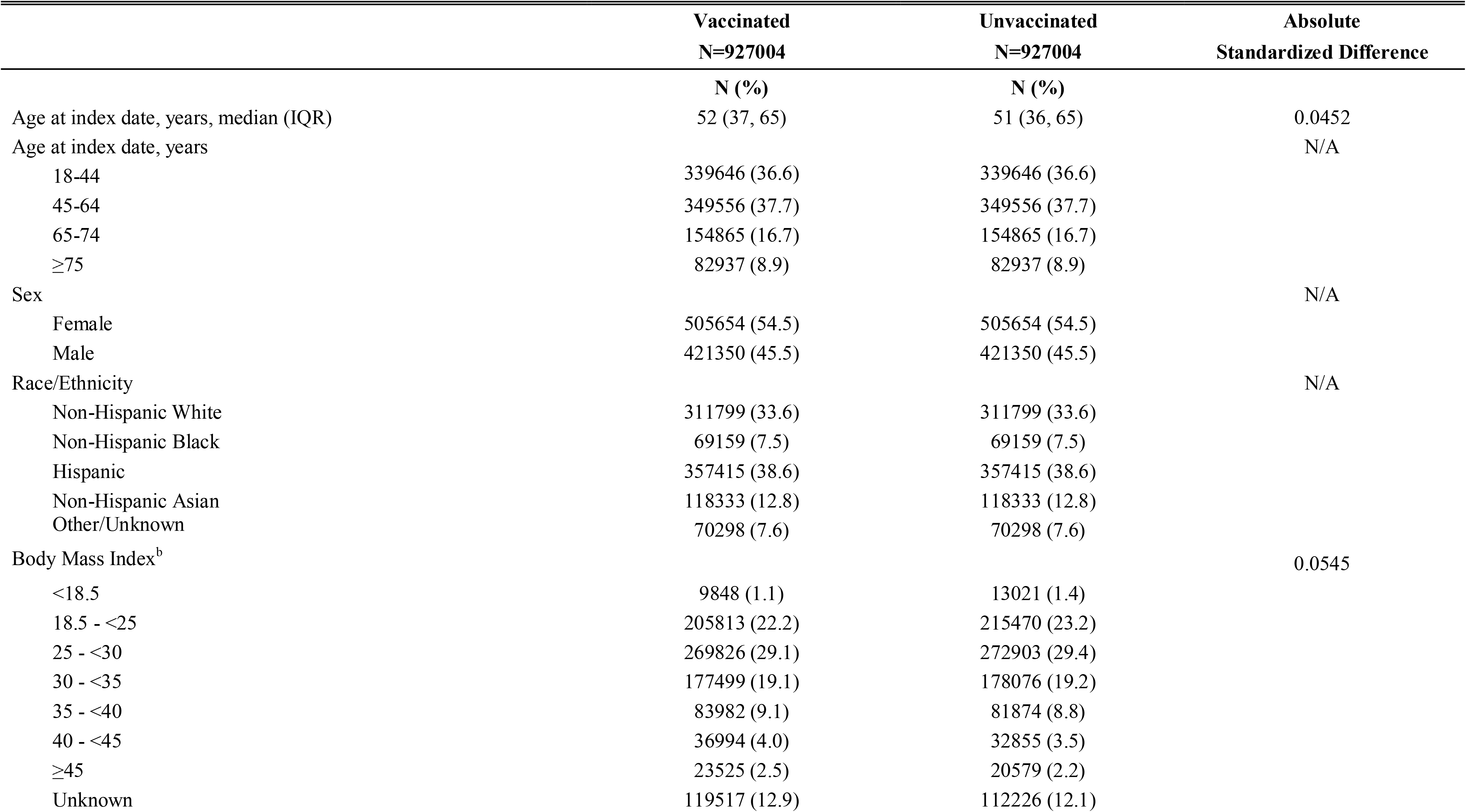
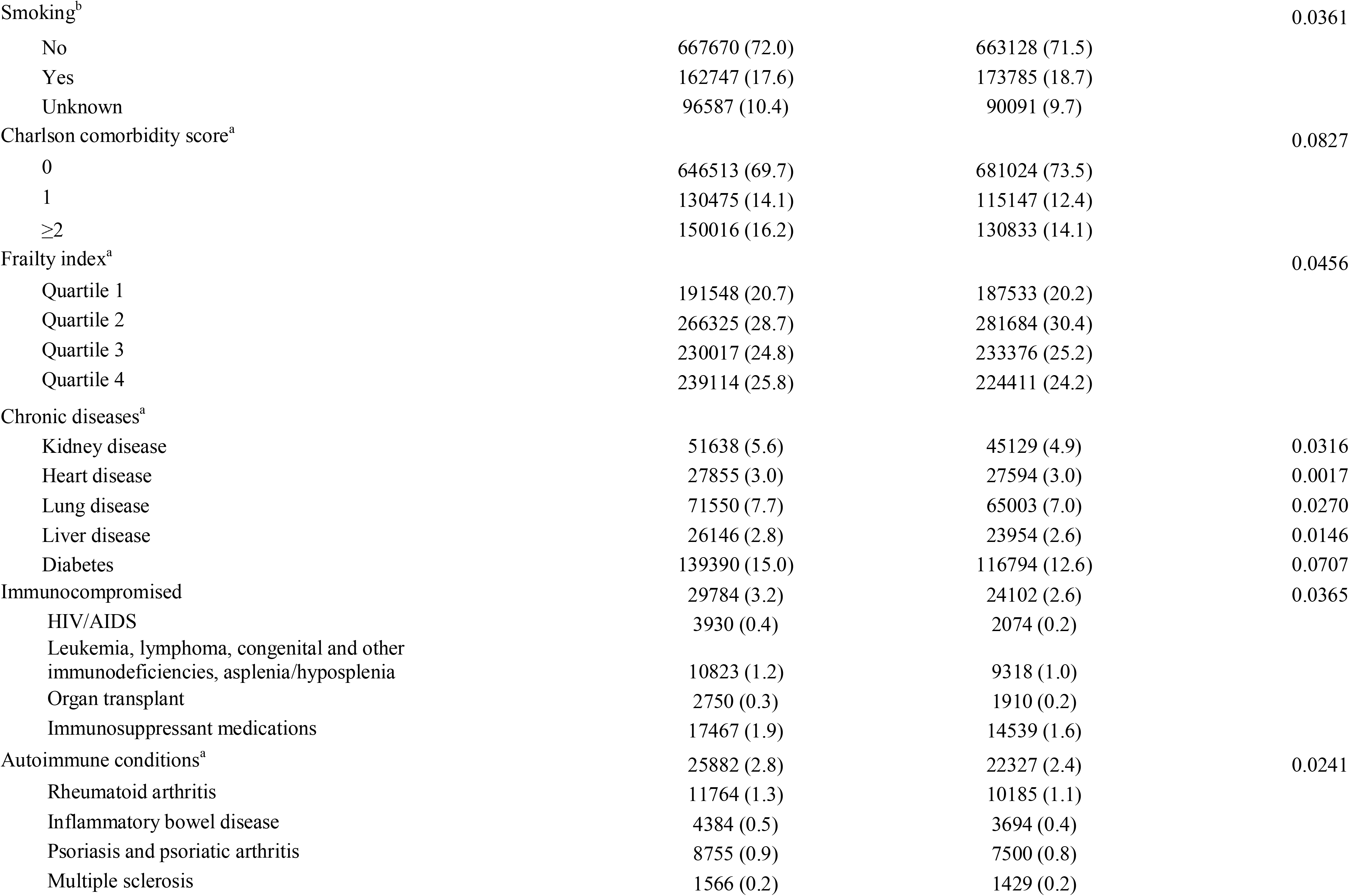
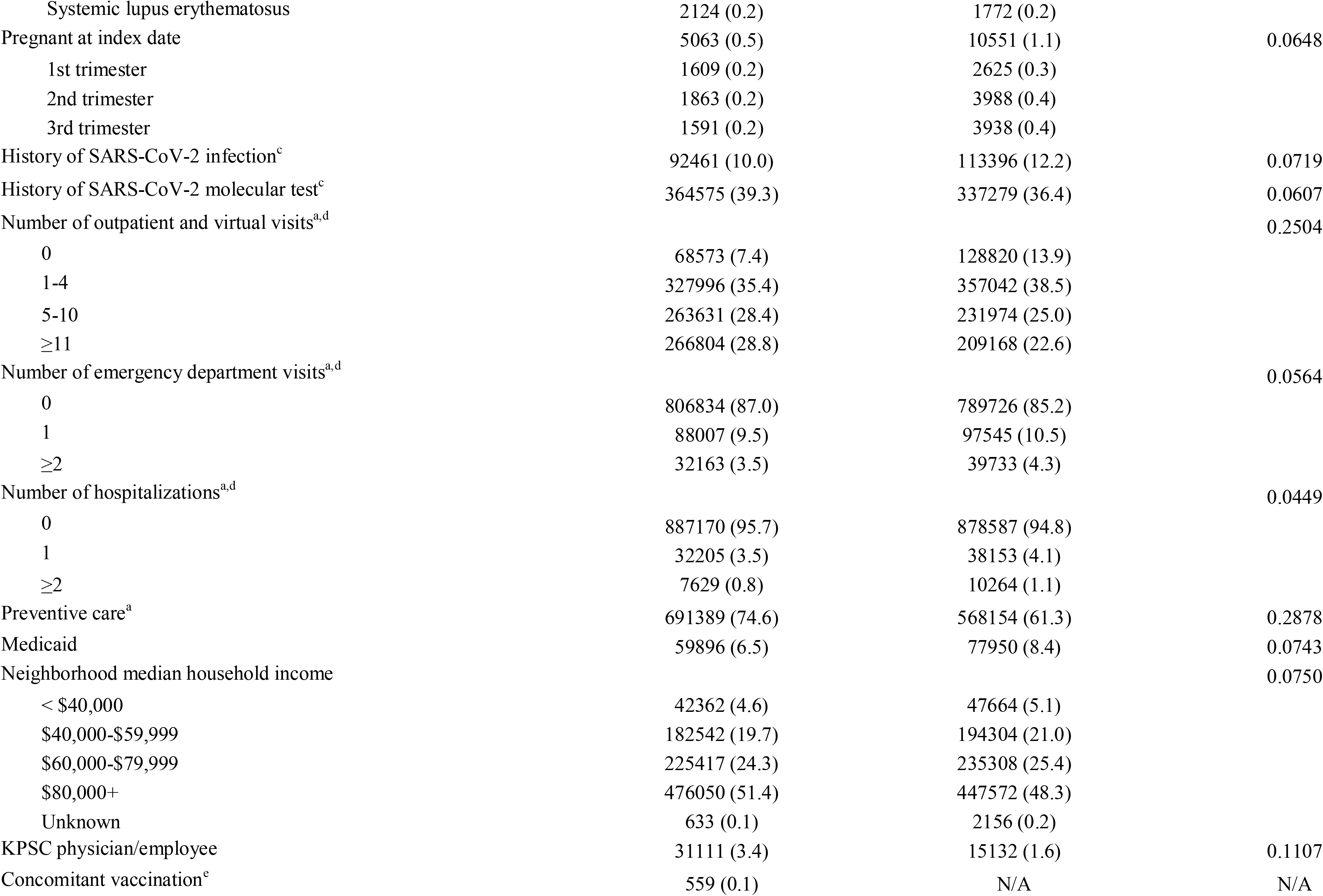
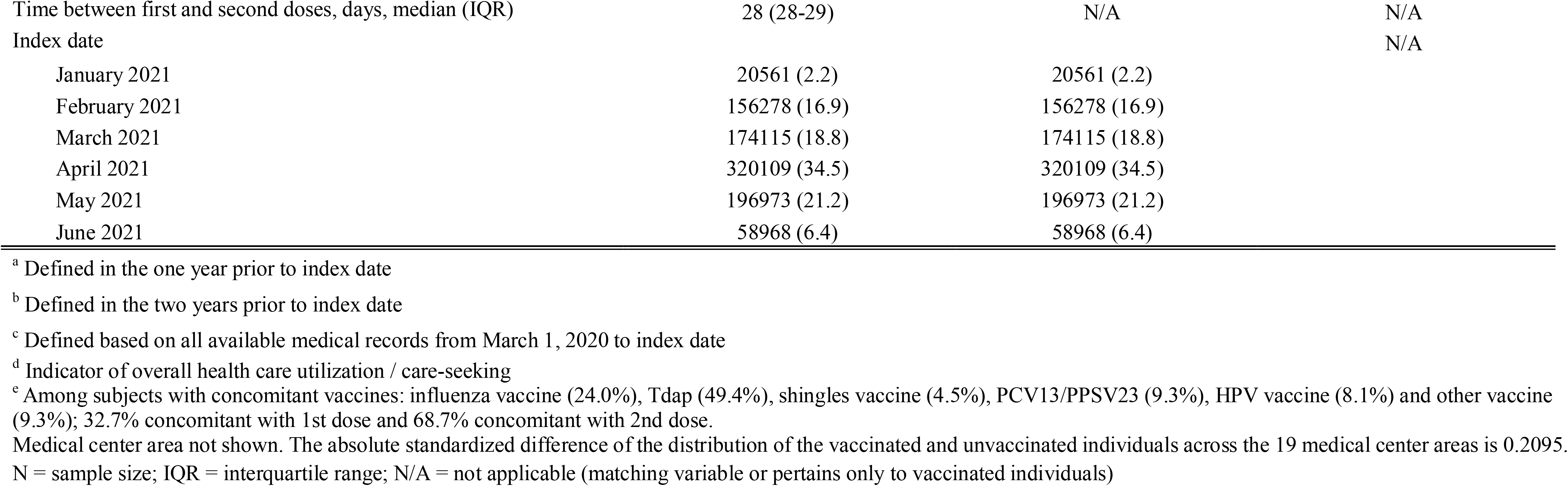
Baseline characteristics of 2-dose mRNA-1273 vaccinated and unvaccinated cohort

**Figure 1:**
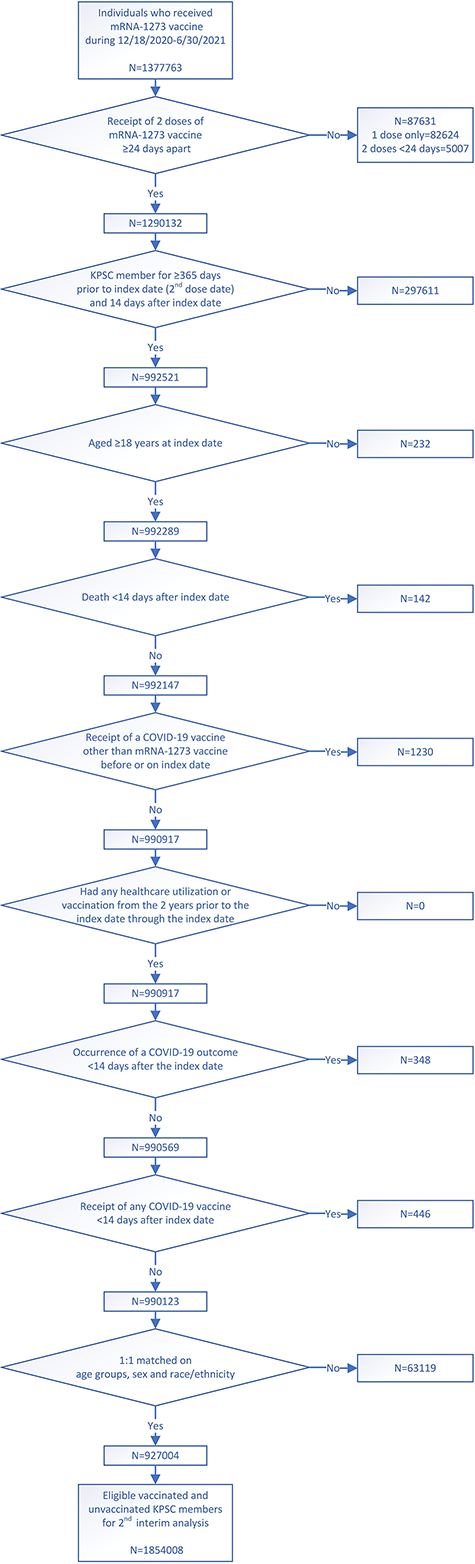
Flow chart for 2-dose mRNA-1273 analysis cohort

### VE against SARS-CoV-2 infection, and COVID-19 hospitalization and hospital death

There were 7,685 cases of SARS-CoV-2 infection among the vaccinated individuals and 16,809 cases among the unvaccinated individuals, with IRs per 1,000 person-years of 19.09 (95% CI: 18.67-19.52) and 94.02 (95% CI: 92.61-95.45), respectively (**Table 2**). IRs per 1,000 person-years for COVID-19 hospitalization and COVID-19 hospital death were 0.60 (95% CI: 0.53-0.69) and 0.03 (95% CI: 0.02-0.06), respectively, among vaccinated individuals and 13.07 (95% CI: 12.55-13.61) and 0.90 (95% CI: 0.77-1.05), respectively, among the unvaccinated individuals. The cumulative incidences of SARS-CoV-2 infection, and COVID-19 hospitalization and hospital death were all significantly higher in unvaccinated individuals than vaccinated individuals (log-rank test p<0.001) (**Figures 2a-c**). The adjusted VE (**Table 2**) was 82.8% (95% CI: 82.2-83.3%) against SARS-CoV-2 infection; 96.1% (95% CI: 95.5-96.6%) against COVID-19 hospitalization; and 97.2% (95% CI: 94.8-98.4%) against COVID-19 hospital death.

**Table 2.** Incidence rates, hazard ratios, and vaccine effectiveness of 2 doses of mRNA-1273 in preventing SARS-CoV-2 infection, and COVID-19 hospitalization and hospital death

| Outcomes | Vaccinated<br>(N=927004) |  | Unvaccinated<br>(N=927004) |  | Hazard Ratio (95% CI) |  | VE % (95% CI) |  |
| --- | --- | --- | --- | --- | --- | --- | --- | --- |
|  | Number<br>of cases | Incidence per 1000<br>person-years<br>(95% CI) | Number<br>of cases | Incidence per 1000<br>person-years<br>(95% CI) | Unadjusted | Adjusted<sup>a</sup> | Unadjusted | Adjusted<sup>a</sup> |
| SARS-CoV-2 infection | 7685 | 19.09 (18.67-19.52) | 16809 | 94.02 (92.61-95.45) | 0.17 (0.16-0.17) | 0.17 (0.17-0.18) | 83.4 (82.9 -83.9) | 82.8 (82.2-83.3) |
| COVID-19 hospitalization | 244 | 0.60 (0.53-0.69) | 2361 | 13.07 (12.55-13.61) | 0.04 (0.03-0.04) | 0.04 (0.03-0.05) | 96.4 (95.9-96.8) | 96.1 (95.5-96.6) |
| COVID-19 hospital death | 13 | 0.03 (0.02-0.06) | 163 | 0.90 (0.77-1.05) | 0.03 (0.02-0.05) | 0.03 (0.02-0.05) | 96.9 (94.5-98.2) | 97.2 (94.8-98.4) |

<sup>a</sup> Adjusted for covariates age groups (4 categories), sex, race/ethnicity, index date (in months), number of outpatient and virtual visits, preventive care, KPSC physician/employee status, medical center area.
N = sample size; CI = confidence interval; VE = vaccine effectiveness

**Figure 2.**
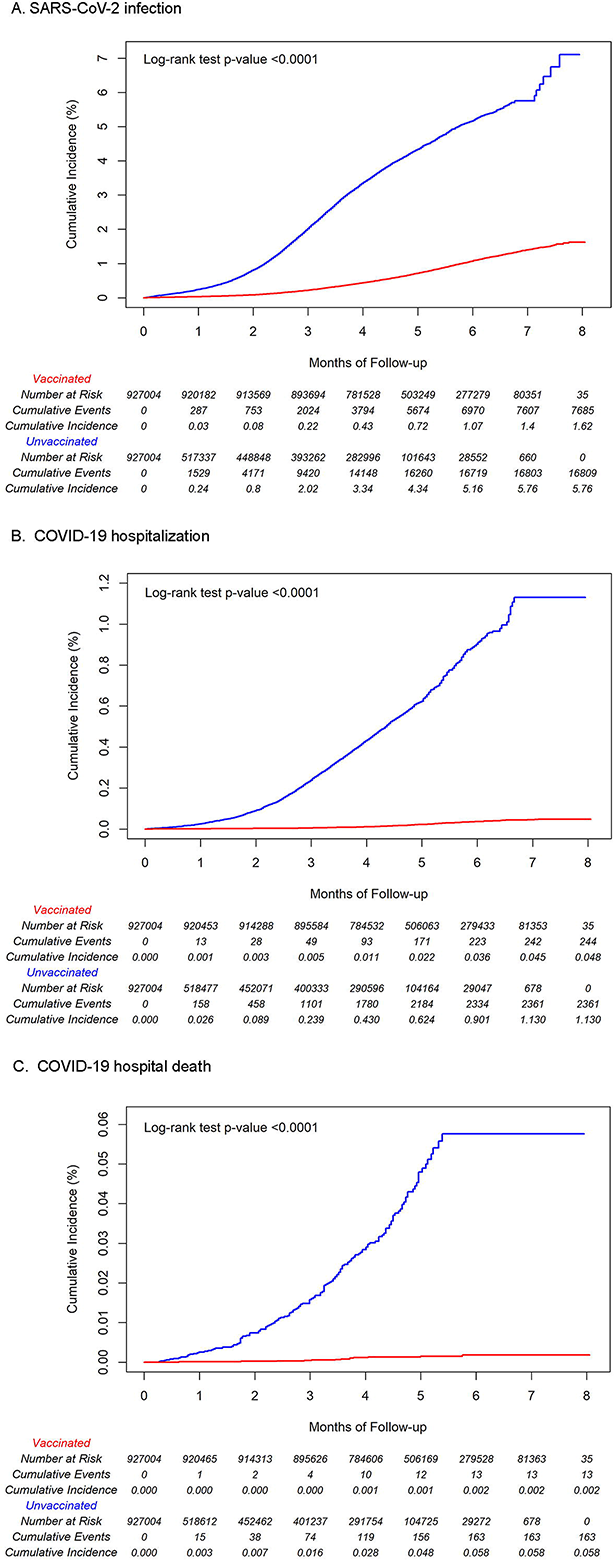
Cumulative incidence estimates by vaccination status in 2-dose mRNA-1273 cohort a. SARS-CoV-2 infection b. COVID-19 hospitalization c. COVID-19 hospital death

### VE against SARS-CoV-2 infection by subgroups

Among vaccinated and unvaccinated groups, the IR of SARS-CoV-2 infection was higher in those □65 years as compared to those ≥65 years (**Table 3**). IRs among vaccinated and unvaccinated females were higher than their male counterparts. IRs were higher among Hispanic individuals in the vaccinated group (23.03 [95% CI: 22.28-23.81]) and among non-Hispanic Black individuals in the unvaccinated group (109.90 [95% CI: 104.47-115.61]) when compared to the other racial/ethnic groups. Despite differences in IRs of SARS-CoV-2 infection across age, sex, and race/ethnicity groups, the adjusted VE against SARS-CoV-2 infection was similar by age, sex, and race/ethnicity, ranging from 77.9% to 87.0%. Among the subgroup of individuals who completed their 2-dose series in June 2021 and their unvaccinated matched counterparts, the IRs for SARS-CoV-2 infection during the Delta period were 20.38 (95% CI: 18.27-22.75) and 146.73 (95% CI: 140.40-153.35), respectively (**Table 3**). Adjusted VE against SARS-CoV-2 infection comparing individuals vaccinated during the Delta period and their unvaccinated counterparts was 86.5% (95% CI: 84.8-88.0%).

**Table 3.** Incidence rate, hazard ratio, and vaccine effectiveness of 2 doses of mRNA-1273 in preventing SARS-CoV-2 infection by age, sex, race/ethnicity, and Delta period subgroups

|  | Vaccinated<br>(N=927004) |  | Unvaccinated<br>(N=927004) |  | Hazard Ratio (95% CI) |  | VE % (95% CI) |  |
| --- | --- | --- | --- | --- | --- | --- | --- | --- |
|  | Number of cases | Incidence per 1000 person-years (95% CI) | Number of cases | Incidence per 1000 person-years (95% CI) | Unadjusted | Adjusted<sup>a</sup> | Unadjusted | Adjusted<sup>a</sup> |
| Age at index date, years |  |  |  |  |  |  |  |  |
| 18-44 | 3196 | 23.05 (22.26-23.86) | 7145 | 109.90 (107.38-112.48) | 0.16 (0.15-0.17) | 0.17 (0.17-0.18) | 84.0 (83.3 -84.7) | 82.6 (81.7 -83.5) |
| 45-64 | 2811 | 19.15 (18.46-19.88) | 6912 | 104.79 (102.35-107.29) | 0.14 (0.13-0.15) | 0.15 (0.15-0.16) | 85.9 (85.2 -86.5) | 84.6 (83.8 -85.4) |
| 65-74 | 1098 | 14.64 (13.80-15.53) | 1877 | 62.57 (59.81-65.47) | 0.19 (0.18-0.21) | 0.18 (0.17-0.20) | 80.8 (79.3 -82.2) | 81.6 (79.9 -83.1) |
| ≥75 | 580 | 13.78 (12.71-14.95) | 875 | 49.13 (45.98-52.49) | 0.24 (0.22-0.27) | 0.22 (0.20-0.25) | 75.9 (73.2 -78.3) | 77.9 (75.1 -80.3) |
| Sex |  |  |  |  |  |  |  |  |
| Female | 4459 | 19.97 (19.39-20.56) | 9116 | 96.18 (94.23-98.18) | 0.17 (0.16-0.17) | 0.18 (0.17-0.18) | 83.2 (82.6 -83.8) | 82.4 (81.7 -83.1) |
| Male | 3226 | 18.00 (17.39-18.64) | 7693 | 91.58 (89.56-93.65) | 0.16 (0.16-0.17) | 0.17 (0.16-0.18) | 83.7 (83.0 -84.4) | 83.1 (82.3 -83.9) |
| Race/Ethnicity |  |  |  |  |  |  |  |  |
| Non-Hispanic White | 2601 | 18.88 (18.16-19.62) | 6061 | 94.48 (92.13-96.89) | 0.17 (0.16-0.18) | 0.18 (0.17-0.19) | 83.2 (82.4 -84.0) | 82.1 (81.1 -83.0) |
| Non-Hispanic Black | 523 | 17.41 (15.98-18.97) | 1497 | 109.90 (104.47-115.61) | 0.14 (0.12-0.15) | 0.13 (0.12-0.15) | 86.2 (84.7 -87.5) | 87.0 (85.5 -88.4) |
| Hispanic | 3488 | 23.03 (22.28-23.81) | 7177 | 103.15 (100.79-105.57) | 0.18 (0.17-0.19) | 0.18 (0.17-0.19) | 82.1 (81.3 -82.9) | 81.8(80.9 -82.6) |
| Non-Hispanic Asian | 612 | 11.37 (10.50-12.31) | 940 | 52.47 (49.22-55.93) | 0.16 (0.14-0.18) | 0.17 (0.15-0.19) | 83.9 (82.1 -85.6) | 83.4 (81.2 -85.4) |
| Delta period<sup>b</sup> | 319 | 20.38 (18.27-22.75) | 1973 | 146.73 (140.40-153.35) | 0.14 (0.12-0.15) | 0.13 (0.12-0.15) | 86.3 (84.6 -87.9) | 86.5 (84.8 -88.0) |

<sup>a</sup> Adjusted for covariates age groups (in 4 categories, and not adjusted in the age subcohort analysis), sex (not adjusted in the sex subcohort analysis), race/ethnicity (not adjusted in the race/ethnicity subcohort analysis), index date (in months), number of outpatient and virtual visits, preventive care, KPSC physician/employee status, medical center area.
<sup>b</sup> Individuals with index dates in June 2021 were followed up to September 30, 2021
N = sample size; CI = confidence interval; VE = vaccine effectiveness

### Durability of protection

Adjusted VE against SARS-CoV-2 infection and COVID-19 hospitalization by months after vaccination both overall and by age groups are presented in **Supplementary Table 1**. Adjusted VE against SARS-CoV-2 infection decreased from 88.0% at 0-<2 months to 75.5% at 6-<8 months (**Figure 3**). Adjusted VE against COVID-19 hospitalization remained stable, from 95.9% in 0-<2 months to 94.5% in 6-<8 months. The durability of protection VE estimates by age groups for both SARS-CoV-2 infection and COVID-19 hospitalization were similar to the overall results.

**Figure 3.**
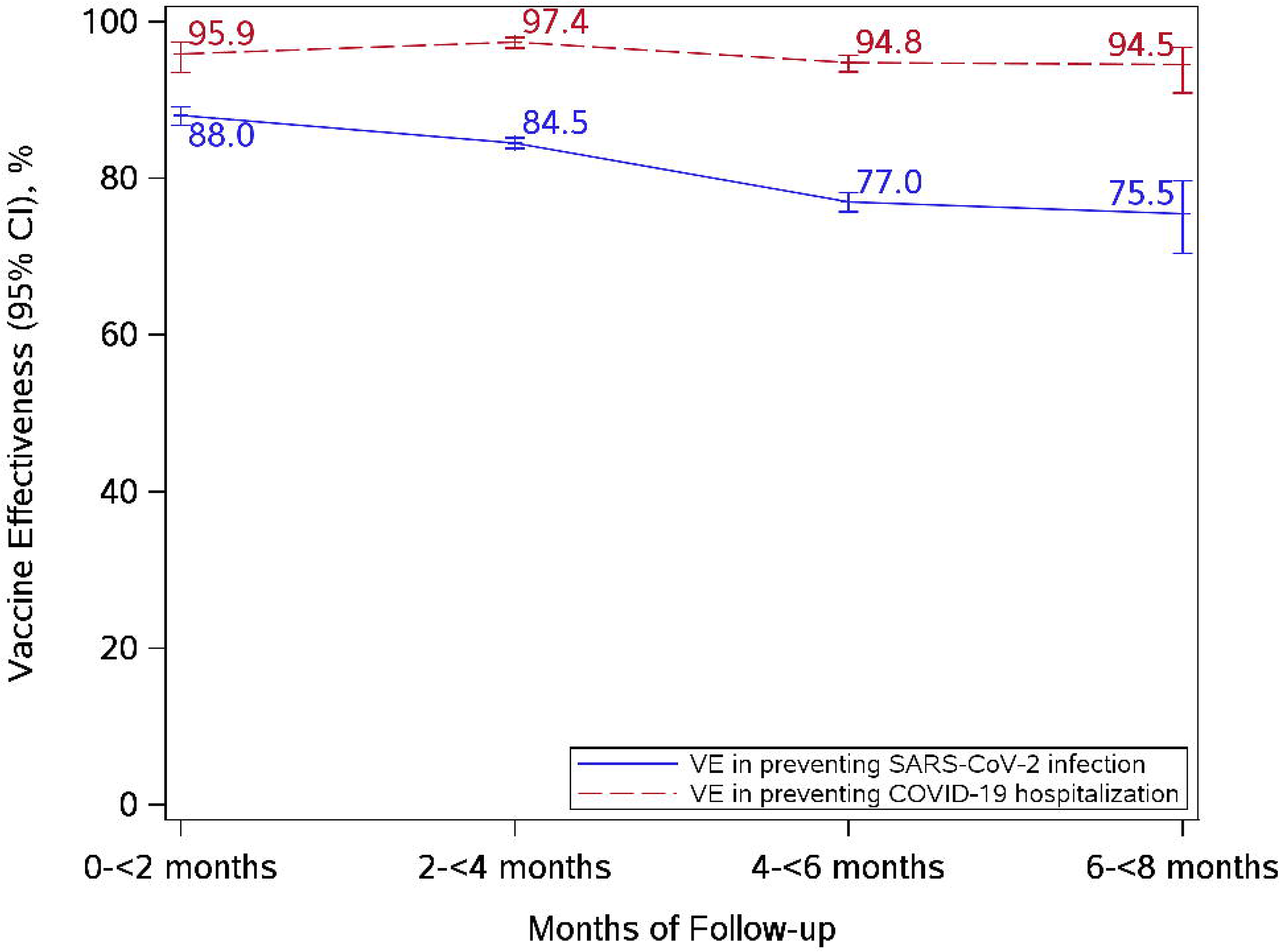
Vaccine effectiveness of 2 doses of mRNA-1273 in preventing SARS-CoV-2 infection and COVID-19 hospitalization by months after vaccination

## Discussion

This interim analysis was conducted among a large cohort to assess the effectiveness and durability of 2 doses of mRNA-1273 under real-world conditions. The study results confirmed the high effectiveness of 2 doses of mRNA-1273, with VE against SARS-CoV-2 infection of 82.8%, VE against COVID-19 hospitalization of 96.1%, and VE against COVID-19 hospital death of 97.2%. Furthermore, VE against COVID-19 hospitalization remained robust and stable over 8 months of follow-up.

Moderate waning was observed for VE against SARS-CoV-2 infection, with a VE of 75.5% at 6-<8 months post-vaccination. The individuals included in the cohort were vaccinated through June 2021 and then followed through September 2021, overlapping with the rapid spread of the Delta variant in the US. Additionally, for individuals who completed the two-dose series in June 2021, VE against SARS-CoV-2 infection during the Delta period remained high at 86.5% through the end of September 2021.

These results add to a growing body of work evaluating the VE of mRNA-1273 in the general population. A case-control study across 21 hospitals in 18 US states assessing the VE of mRNA-1273 in preventing COVID-19 hospitalizations during March 11, 2021 to August 15, 2021 reported a VE of 93% (95% CI: 91%-95%), similar to the VE found in this study [25]. The findings of our study are also consistent with and extend those of our prior interim analysis [1] with follow-up through June 2021. VE against COVID-19 hospitalization and hospital death was higher than the VE against SARS-CoV-2 infection, corroborating previous evidence that mRNA vaccines offer greater protection against more severe disease [1, 26, 27].

We previously found mRNA-1273 to have a high VE against SARS-CoV-2 infection before the Delta variant spread in the US [1]. The Delta variant started emerging in the US in late May 2021 and quickly became the dominant variant during June through August 2021 [28]. A study in Qatar evaluated VE of mRNA-1273 against the Delta variant [29], and found a VE against SARS-CoV-2 infection of 73.1% (95% CI: 67.5%-77.8%). In the current study, VE against SARS-CoV-2 infection was evaluated for the Delta period in a subset of individuals newly vaccinated during June 2021 and followed up through September 2021; VE against Delta after three months of follow-up was 86.5%, similar to the VE observed in our prior interim analysis (87.4%) [1]. This finding is less subject to waning and suggests that mRNA-1273 generates protection against the Delta variant.

The current study is also one of the first to examine the durability of the 2-dose mRNA-1273 vaccine. While VE against COVID-19 hospitalization was stable during the follow-up, VE against SARS-CoV-2 infection steadily decreased from 88.0% to 75.5% across the 8 months of follow-up. The same VE waning effects were apparent across age categories (18-64 years and ≥65 years). These findings are consistent with the findings of our previous study that used a test-negative design [13]. Due to this steady decrease in VE, our findings support the current mRNA-1273 booster recommendations of receiving a booster shot at least 6 months after completing the primary series [30].

One strength of the current study is the matched cohort design which allows for generalizability to the general population eligible for mRNA-1273. Another strength is KPSC’s diverse member population. The study also utilized KPSC’s EHR and was able to gather comprehensive data on millions of members; the data included COVID-19 vaccine information, demographics, medical history, and health care utilization. However, residual confounding from unmeasured factors might have still been present. Some health seeking behaviors such as adherence to masking guidelines and occupation are not captured by the EHR; these may contribute to differences in COVID-19 risk and testing behaviors between vaccinated and unvaccinated individuals. Misclassification of SARS-CoV-2 infection from false positive or false negative test results or from inaccurate diagnosis codes from outside claims may have been another limitation. This non-differential misclassification could have underestimated the VE. Lastly, individuals vaccinated during the Delta period were younger as vaccines had recently become available to the general population and may have had fewer COVID-19 risk factors, which could have led to overestimation of VE during the Delta period.

In conclusion, this second interim analysis of an ongoing cohort study found that VE of 2 doses of mRNA-1273 against SARS-CoV-2 infection declined moderately over the course of 8 months, but VE against COVID-19 hospitalization remained robust and stable over the same period. In addition, VE against SARS-CoV-2 infection in newly vaccinated individuals during the Delta period remained high. Continued long-term follow-up is needed to fully evaluate the real-world effectiveness of mRNA-1273 overall and in different subgroups of the population over time.

## Data Availability

Individual-level data reported in this study are not publicly shared. Upon request, and subject to review, KPSC may provide the deidentified aggregate-level data that support the findings of this study. Deidentified data (including participant data as applicable) may be shared upon approval of an analysis proposal and a signed data access agreement

## Author contributions

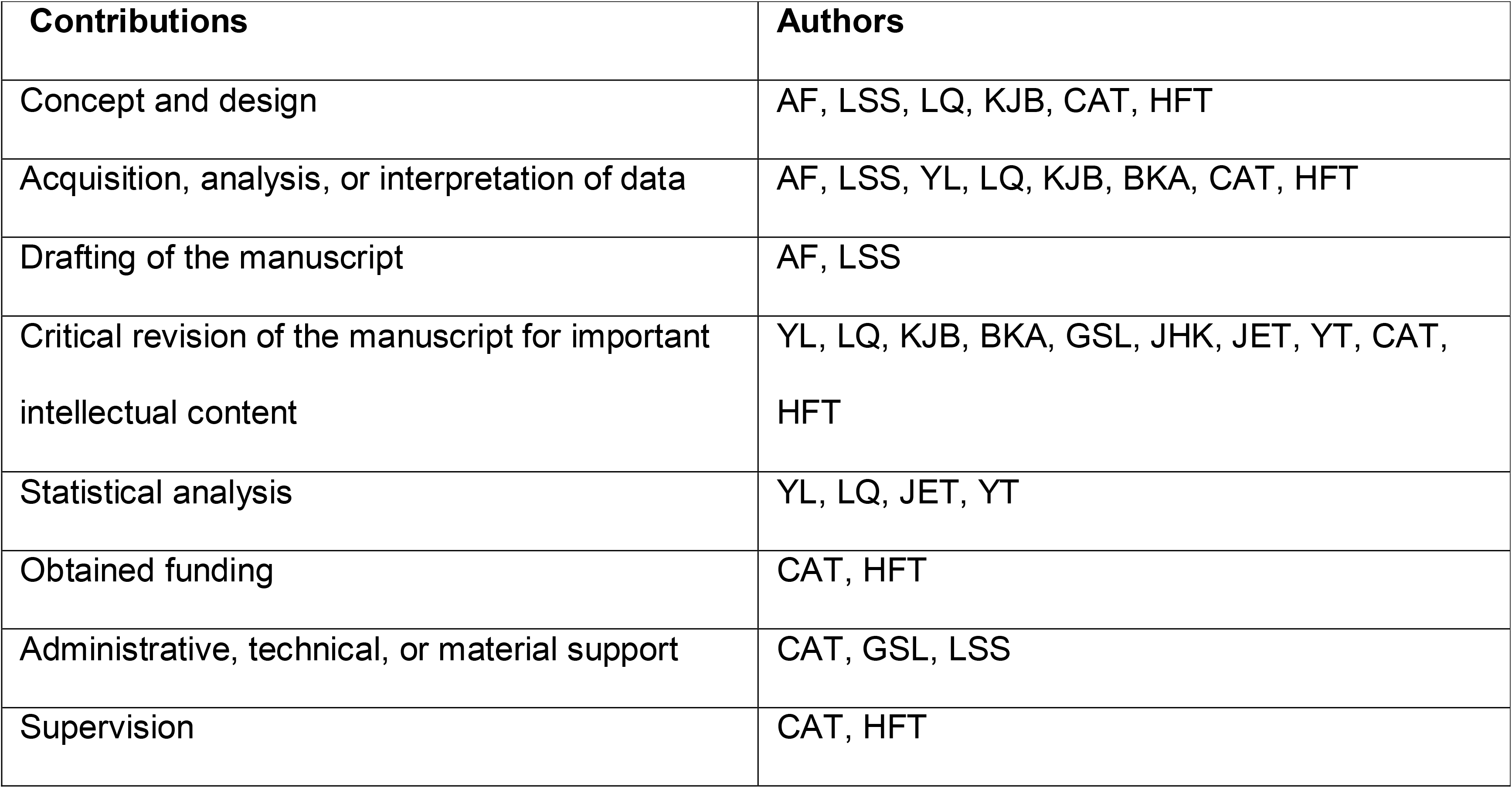

## Declaration of interests

AF, LSS, YL, LQ, BKA, GSL, JHK, JET, YT, and HFT are employees of Kaiser Permanente Southern California, which has been contracted by Moderna to conduct this study. KJB is an adjunct investigator at Kaiser Permanente Southern California. CAT is an employee of and a shareholder in Moderna Inc. AF received funding from Pfizer, GlaxoSmithKline, and Gilead unrelated to this manuscript. LSS received funding from GlaxoSmithKline, Dynavax, and Seqirus unrelated to this manuscript. YL received funding from GlaxoSmithKline, Seqirus and Pfizer unrelated to this manuscript. LQ received funding from GlaxoSmithKline and Dynavax unrelated to this manuscript. KJB received funding from GlaxoSmithKline, Dynavax, Pfizer, Gilead, and Seqirus unrelated to this manuscript. BKA received funding from GlaxoSmithKline, Dynavax, Seqirus and Pfizer unrelated to this manuscript. GSL received funding from GlaxoSmithKline unrelated to this manuscript. JHK received funding from GlaxoSmithKline unrelated to this manuscript. JET received funding from Pfizer unrelated to this manuscript. YT received funding from GlaxoSmithKline unrelated to this manuscript. HFT received funding from GlaxoSmithKline and Seqirus unrelated to this manuscript; HFT also served in advisory boards for Janssen and Pfizer.

## Funding

This work was supported by Moderna Inc.

## Acknowledgments

This study was funded by Moderna Inc. The authors would like to acknowledge the following Kaiser Permanente Southern California staff: Radha Bathala, Maria Navarro, Elsa Olvera, Brittany Brown, Jeannie Song, and Stephanie Reese Rillon for their contributions in manual chart review; Harpreet Takhar, Michael Aragones, Soon Kyu Choi, Jennifer Charter, Julie Stern, Joy Gelfond, and Lee Childs for their coordination in processing SARS-CoV-2 specimens; and Raul Calderon, Kourtney Kottman, Ana Acevedo, Elmer Ayala, and Jonathan Arguello for their technical and laboratory support in processing SARS-CoV-2 specimens. The authors would like to acknowledge Helix OpCo, LLC, for their whole genome sequencing of SARS-CoV-2 specimens. The authors would also like to acknowledge the contributions by Moderna staff: Yamuna Paila, PhD, and Julie Vanas. Medical writing and editorial assistance were provided by Srividya Ramachandran, PhD, and Jared Mackenzie, PhD, of MEDiSTRAVA in accordance with Good Publication Practice (GPP3) guidelines, funded by Moderna Inc, and under the direction of the authors. The authors thank the patients of Kaiser Permanente for their partnership with us to improve their health. Their information, collected through our electronic health record systems, leads to findings that help us improve care for our members and can be shared with the larger community.

## Ethical Approval

The study was reviewed and approved by the KPSC Institutional Review Board (IRB #12758). A waiver of informed consent was obtained as this is an observational study of the authorized and recommended Moderna COVID-19 vaccine administered during routine clinical care. To facilitate the conduct of this study, a waiver was obtained for written HIPAA authorization for research involving use of the EHR.

## Notes

### Summary of Updates

There have been no updates to the manuscript file. The supplementary information has been included in this revision.

