## Supplementary Information for "Durability of mRNA-1273 against COVID-19 in the time of Delta: Interim results from an observational cohort study"

**Supplementary Materials**

#### **Supplementary Table 1**. Incidence rate, hazard ratio, and vaccine effectiveness of 2 doses of mRNA-1273 vaccine in preventing SARS-CoV-2 infection and COVID-19 hospitalization, stratified by age, by months after vaccination

|  | **Vaccinated** | | | **Unvaccinated** | | | **Hazard Ratio (95% CI)** | | **VE % (95% CI)** | |
| --- | --- | --- | --- | --- | --- | --- | --- | --- | --- | --- |
| **Outcomes** | **N** | **Number of cases** | **Incidence per 1000 person-years  (95% CI)** | **N** | **Number of cases** | **Incidence per 1000 person-years (95% CI)** | **Unadjusted** | **Adjusted^a^** | **Unadjusted** | **Adjusted^a^** |
| SARS-CoV-2 infection |  |  |  |  |  |  |  |  |  |  |
| 0-<2 months | 927004 | 528 | 4.36 (4.00-4.75) | 927004 | 2786 | 36.14 (34.82-37.51) | 0.12 (0.11-0.13) | 0.12 (0.11-0.13) | 88.4 (87.3-89.4) | 88.0 (86.8-89.1) |
| 2-<4 months | 916323 | 2556 | 17.02 (16.37-17.69) | 469386 | 9813 | 141.55 (138.78-144.38) | 0.12 (0.11-0.12) | 0.16 (0.15-0.16) | 88.2 (87.6-88.7) | 84.5 (83.8-85.2) |
| 4-<6 months | 843804 | 3455 | 33.46 (32.36-34.60) | 336656 | 4022 | 132.60 (128.57-136.77) | 0.25 (0.24-0.26) | 0.23 (0.22-0.24) | 75.1 (73.9-76.2) | 77.0 (75.7-78.2) |
| 6-<8 months | 326241 | 1141 | 41.11 (38.80-43.57) | 40040 | 188 | 92.30 (80.01-106.49) | 0.46 (0.39-0.54) | 0.25 (0.20-0.30) | 53.8 (46.0-60.5) | 75.5 (70.4-79.7) |
| Aged 18-64 years old |  |  |  |  |  |  |  |  |  |  |
| 0-<2 months | 689202 | 418 | 4.65 (4.22-5.11) | 689202 | 2373 | 39.86 (38.29-41.50) | 0.11 (0.10-0.12) | 0.12 (0.10-0.13) | 88.9 (87.7-90.0) | 88.4 (87.1-89.6) |
| 2-<4 months | 680159 | 2293 | 20.65 (19.82-21.51) | 361409 | 8968 | 169.98 (166.50-173.53) | 0.12 (0.11-0.13) | 0.15 (0.15-0.16) | 88.1 (87.5-88.6) | 84.6 (83.9-85.4) |
| 4-<6 months | 613565 | 2481 | 36.36 (34.96-37.82) | 247334 | 2694 | 145.35 (139.96-150.94) | 0.24 (0.23-0.26) | 0.22 (0.21-0.24) | 75.9 (74.5-77.2) | 77.6 (76.0-79.0) |
| 6-<8 months | 164711 | 811 | 50.69 (47.32-54.30) | 2386 | 22 | 152.32 (100.29-231.33) | 0.33 (0.22-0.51) | 0.28 (0.18-0.45) | 66.6 (49.0-78.2) | 71.6 (54.7-82.2) |
| Aged ≥65 years old | |  |  |  |  |  |  |  |  |  |
| 0-<2 months | 237802 | 110 | 3.53 (2.93-4.26) | 237802 | 413 | 23.53 (21.36-25.91) | 0.15 (0.12-0.19) | 0.13 (0.11-0.17) | 84.7 (81.1-87.6) | 86.5 (83.2-89.2) |
| 2-<4 months | 236164 | 263 | 6.71 (5.95-7.58) | 107977 | 845 | 51.01 (47.68-54.57) | 0.13 (0.11-0.15) | 0.16 (0.14-0.18) | 87.1 (85.2-88.8) | 84.2 (81.7-86.3) |
| 4-<6 months | 230239 | 974 | 27.81 (26.12-29.61) | 89322 | 1328 | 112.58 (106.69-118.80) | 0.24 (0.22-0.27) | 0.23 (0.21-0.26) | 75.6 (73.5-77.5) | 76.8 (74.5-78.8) |
| 6-<8 months | 161530 | 330 | 28.08 (25.21-31.28) | 37654 | 166 | 87.72 (75.34-102.14) | 0.33 (0.27-0.39) | 0.22 (0.18-0.28) | 67.4 (60.5-73.0) | 77.7 (72.3-82.1) |
| COVID-19 hospitalization |  |  |  |  |  |  |  |  |  |  |
| 0-<2 months | 927004 | 21 | 0.17 (0.11-0.27) | 927004 | 301 | 3.90 (3.48-4.36) | 0.04 (0.03-0.07) | 0.04 (0.03-0.06) | 95.8 (93.5-97.3) | 95.9 (93.5-97.4) |
| 2-<4 months | 916826 | 55 | 0.37 (0.28-0.48) | 471487 | 1226 | 17.46 (16.51-18.46) | 0.02 (0.02-0.03) | 0.03 (0.02-0.03) | 97.9 (97.3-98.4) | 97.4 (96.6-98.0) |
| 4-<6 months | 846468 | 132 | 1.27 (1.07-1.51) | 344860 | 771 | 24.77 (23.08-26.58) | 0.05 (0.04-0.06) | 0.05 (0.04-0.06) | 95.1 (94.1-95.9) | 94.8 (93.6-95.7) |
| 6-<8 months | 328421 | 36 | 1.29 (0.93-1.78) | 40715 | 63 | 30.41 (23.76-38.93) | 0.05 (0.03-0.07) | 0.06 (0.03-0.09) | 95.3 (92.8-96.9) | 94.5 (90.9-96.7) |
| Aged 18-64 years old |  |  |  |  |  |  |  |  |  |  |
| 0-<2 months | 689202 | 11 | 0.12 (0.07-0.22) | 689202 | 195 | 3.27 (2.84-3.76) | 0.03 (0.02-0.06) | 0.04 (0.02-0.07) | 96.5 (93.6-98.1) | 96.2 (93.0-98.0) |
| 2-<4 months | 680563 | 30 | 0.27 (0.19-0.39) | 363280 | 957 | 17.86 (16.76-19.03) | 0.01 (0.01-0.02) | 0.02 (0.01-0.03) | 98.5 (97.9-99.0) | 98.1 (97.3-98.7) |
| 4-<6 months | 615914 | 35 | 0.51 (0.37-0.71) | 254857 | 306 | 15.95 (14.26-17.84) | 0.03 (0.02-0.05) | 0.03 (0.02-0.05) | 96.5 (95.0-97.6) | 96.6 (95.0-97.8) |
| 6-<8 months | 166067 | 8 | 0.49 (0.25-0.99) | 2460 | 2 | 13.48 (3.37-53.91) | 0.04 (0.01-0.18) | 0.01 (0.00-0.21) | 96.2 (82.0-99.2) | 98.7 (78.6-99.9) |
| Aged ≥65 years old | |  |  |  |  |  |  |  |  |  |
| 0-<2 months | 237802 | 10 | 0.32 (0.17-0.60) | 237802 | 106 | 6.03 (4.99-7.30) | 0.05 (0.03-0.10) | 0.05 (0.02-0.09) | 94.8 (90.0-97.3) | 95.2 (90.6-97.5) |
| 2-<4 months | 236263 | 25 | 0.64 (0.43-0.94) | 108207 | 269 | 16.17 (14.35-18.23) | 0.04 (0.03-0.06) | 0.05 (0.03-0.08) | 96.1 (94.2-97.4) | 95.0 (92.4-96.7) |
| 4-<6 months | 230554 | 97 | 2.76 (2.26-3.37) | 90003 | 465 | 38.96 (35.58-42.67) | 0.07 (0.06-0.09) | 0.07 (0.05-0.09) | 93.0 (91.2-94.3) | 93.2 (91.4-94.6) |
| 6-<8 months | 162354 | 28 | 2.37 (1.63-3.43) | 38255 | 61 | 31.71 (24.68-40.76) | 0.08 (0.05-0.12) | 0.06 (0.04-0.10) | 92.4 (88.0-95.2) | 93.8 (89.8-96.3) |

^a^ Adjusted for covariates age, sex, race/ethnicity, index date (in months), number of outpatient and virtual visits, preventive care, KPSC physician/employee status, medical center area.

N = sample size; CI = confidence interval; VE = vaccine effectiveness

#### **Study protocol**

Real-World Study of the Effectiveness of Moderna COVID-19 Vaccine

**mRNA-1273-P901 RWS**

**FINAL**

September 7, 2021

Kaiser Permanente Southern California

Hung Fu Tseng, PhD, MPH

Research Scientist III

Katia Bruxvoort, PhD, MPH

Post-Doctoral Research Fellow

Moderna, Inc.

Carla Talarico, PhD, MPH Lead Epidemiologist

**Approvals**

| **Approvers, Organization & Role** | **Approval Status & Signature** | **Date** |
| --- | --- | --- |
| Hung-Fu Tseng  KPSC Research and Evaluation Principal Investigator | [*Hung Fu Tseng*](https://kaiserpermanente.na2.adobesign.com/verifier?tx=CBJCHBCAABAAV2f-jpXBF0LmXHT676YxB_NLGkolFB8y)  [Hung Fu Tseng (Sep 20, 2021 20:31 PDT)](https://kaiserpermanente.na2.adobesign.com/verifier?tx=CBJCHBCAABAAV2f-jpXBF0LmXHT676YxB_NLGkolFB8y) | Sep 20, 2021 |
| Jacqueline Miller  Therapeutic Area Head, Infectious Disease Development  Moderna Inc. | [*Jacqueline M. Miller, MD*](https://kaiserpermanente.na2.adobesign.com/verifier?tx=CBJCHBCAABAAV2f-jpXBF0LmXHT676YxB_NLGkolFB8y)  [Jacqueline M. Miller, MD (Sep 21, 2021 10:42 EDT)](https://kaiserpermanente.na2.adobesign.com/verifier?tx=CBJCHBCAABAAV2f-jpXBF0LmXHT676YxB_NLGkolFB8y) | Sep 21, 2021 |

**Version History**

| **Version** | **Date** | **Primary Author/Reviewer** | **Description** |
| --- | --- | --- | --- |
| 0.1 | January 22, 2021 | Katia Bruxvoort | 1st Draft Submitted to Moderna for review |
| 0.2 | February 9, 2021 | Katia Bruxvoort | 2nd Draft Submitted to Moderna for review |
| 1.0 | February 17, 2021 | Katia Bruxvoort | Final Draft Submitted to Moderna for review |
| 1.1 | May 21, 2021 | Katia Bruxvoort | Draft Submitted to Moderna for review – Update: Addition of adolescents (12-17 years); addition of timing of select secondary analyses; clarifications in response to questions from EMA and CBER. |
| 2.0 | June 4, 2021 | Katia Bruxvoort | Final Draft Submitted to Moderna for review |
| 2.1 | August 4, 2021 | Katia Bruxvoort | Draft Submitted to Moderna for review – Update: Addition of vaccine effectiveness against SARS-CoV-2 variants. |
| 3.0 | September 7, 2021 | Katia Bruxvoort | Final Draft Submitted to Moderna for review |

Table of Contents

List of Tables	5

List of Figures	6

List of Abbreviations	7

1 Synopsis	8
2 Background	10
3 Research Objectives	11
  3.1 Primary Objectives	11
  3.2 Secondary Objectives	11
4 Study Setting	13
  4.1 COVID-19 Vaccination at KPSC	14
  4.2 COVID-19 at KPSC	15
  4.3 SARS-CoV-2 Whole Genome Sequencing	17
5 Study Design Overview	18
6 Study Population	19
  6.1 Eligibility Criteria	19
  6.2 Index Dates	19
7 Measures	21
  7.1 Exposure Definition	21
  7.2 Outcome Definitions	21
  7.3 Other Variable Definitions	22
8 Statistical Analysis for Study Objectives 1-15	25
  8.1 Analysis Plan	25
  8.2 Descriptive Analysis	27
  8.3 Primary Analyses	27
  8.4 Secondary Analyses	27
9 Sample Size and Power Estimation	30
10 Statistical Analysis Plan for Vaccine Effectiveness Against SARS-CoV-2 Variants (Study Objectives 16-17)	32
   10.1 Test-negative design (TND)	32
   10.2 Cohort design	35
   10.3 Other considerations	35
11 Timelines	37
12 Data Management	38
13 Limitations	39
14 Human Subjects Protection	40
15 References	41

### List of Tables

###### Table 1. Demographic characteristics of Kaiser Permanente Southern California members on January 1, 2021 14

###### Table 2. SARS-CoV-2 molecular tests and diagnoses among members of Kaiser Permanente Southern California during 01/01/2020-12/31/2020 16

###### Table 3. List of possible covariates 22

###### Table 4. Power calculations for 1st interim analysis of Primary Objective 1 for various sample sizes and vaccine effectiveness estimates 30

###### Table 5. Power calculations for 1st interim analysis of Primary Objective 2 for various sample sizes and vaccine effectiveness estimates 31

###### Table 6. Estimated Project Timelines 37

### List of Figures

###### Figure 1. Analysis Plan for Study Objectives 1-15 26

### List of Abbreviations

| ACIP AIDS ASD BMI CAIR CDC CI COPD  COVID-19  Ct EHR EUA HIPAA HR HIV ICD-10 IRB IPTW KPSC NLP MCE mRNA PHI  SARS-CoV-2 TND  U.S.  VE | Advisory Committee on Immunization Practices Acquired Immunodeficiency Syndrome Absolute Standardized Difference  Body Mass Index  California Immunization Registry  Centers for Disease Control and Prevention Confidence Interval  Chronic Obstructive Pulmonary Disease Coronavirus Disease 2019  Cycle Threshold Electronic Health Record  Emergency Use Authorization  Health Insurance Portability and Accountability Act of 1996 (U.S.) Hazard Ratio  Human Immunodeficiency Virus  International Classification of Diseases, 10th Revision Institutional Review Board  Inverse Probability of Treatment Weighting Kaiser Permanente of Southern California Natural Language Processing  Multi-County Entity Messenger RNA  Protected Health Information  Severe Acute Respiratory Syndrome Coronavirus 2 Test-Negative Design  United States  Vaccine Effectiveness |
| --- | --- |
| VOC | Variant of Concern |
| VOI | Variant of Interest |
| WGS | Whole Genome Sequencing |

### 1 Synopsis

Severe acute respiratory syndrome coronavirus 2 (SARS-CoV-2), the virus causing the coronavirus disease 2019 (COVID-19) pandemic, has caused high global morbidity and mortality,[1] with multiple SARS-CoV-2 variants of concern (VOCs) and variants of interest (VOIs) emerging over the course of the pandemic. [2-4] COVID-19 vaccines have been developed at unprecedented speed, including the Moderna mRNA-1273 SARS-CoV-2 vaccine (Moderna COVID-19 vaccine, hereafter), which received Emergency Use Authorization (EUA) in the United States (U.S.) on December 18, 2020.[5]

Moderna COVID-19 vaccine is a lipid nanoparticle encapsulated messenger RNA (mRNA)- based vaccine encoding the prefusion-stabilized spike glycoprotein of SARS-CoV-2. In a phase 3 randomized, observer-blinded, placebo-controlled trial enrolling approximately 30,000 participants, vaccine efficacy of Moderna COVID-19 vaccine was 94.1% and was generally consistent across age, sex, race/ethnicity, and risk status. No safety concerns were identified.[6] However, post-authorization studies are critically needed to determine Moderna COVID-19 vaccine effectiveness and durability in real-world settings among diverse populations.

In this protocol, we describe an observational cohort study at Kaiser Permanente Southern California (KPSC), a large, integrated health care system. The primary objectives are to evaluate the vaccine effectiveness (VE) of 2 doses of Moderna COVID-19 vaccine in preventing COVID-19 diagnosis and severe COVID-19 disease. COVID-19 diagnosis will be defined as a positive SARS-CoV-2 molecular diagnostic test among symptomatic or asymptomatic individuals or a diagnosis code. Severe COVID-19 disease will be defined as COVID-19 hospitalization or mortality. KPSC members meeting inclusion criteria will be considered exposed if they received 2 doses of Moderna COVID-19 vaccine during 2021, and the unexposed comparison group will be a similar unvaccinated population. Individuals will be followed through electronic health records for COVID-19 outcomes through December 2023, with 6 interim analyses and a final analysis being conducted.

For first and second interim analyses, unvaccinated individuals will be randomly selected and n:1 matched to vaccinated individuals by age, sex, and race/ethnicity (if sample size allows), and will be assigned an index date based on the vaccination date of their matched vaccinated individual. For subsequent interim analyses and the final analysis, the number of eligible unvaccinated individuals in the KPSC population may be insufficient for individual matching; if so, frequency matching will be employed. In all analyses, efforts will be dedicated to identifying and adjusting for potential confounders. Cox proportional hazards regression will be used to estimate unadjusted and adjusted hazards ratios (HRs). VE (%) will be estimated as (1 – adjusted HR) × 100. Propensity score analyses with inverse probability of treatment weighting (IPTW) will be considered for final analyses.

Secondary analyses will be conducted to examine VE of 2 doses of Moderna COVID-19 vaccine in preventing COVID-19 diagnosis stratified by age, sex, and race/ethnicity, as well as among individuals with chronic conditions, immunocompromised individuals, individuals with autoimmune conditions, frail individuals, pregnant women, and individuals with a history of COVID-19 diagnosis. In addition, we will examine the VE of 2 doses of Moderna COVID-19 vaccine in preventing COVID-19 diagnosis when given concomitantly with another vaccine. We will examine the VE of 2 doses of Moderna COVID-19 vaccine in preventing asymptomatic

COVID-19 and the VE of 2 doses of Moderna COVID-19 vaccine in preventing symptomatic COVID-19. Also, we will examine durability of 2 doses of Moderna COVID-19 vaccine in preventing COVID-19 diagnosis and severe COVID-19, as well as the VE of 1 dose of Moderna COVID-19 vaccine in preventing COVID-19 diagnosis and severe COVID-19. Using a test- negative and cohort study design, we will examine the VE of 2 doses of Moderna COVID-19 vaccine against SARS-CoV-2 variants, and the VE of 1 dose of Moderna COVID-19 vaccine against SARS-CoV-2 variants. Some secondary analyses may be underpowered; their meaningfulness will depend on uptake of Moderna COVID-19 vaccine in these patient populations.

### 2 Background

Severe acute respiratory syndrome coronavirus 2 (SARS-CoV-2), the virus causing the coronavirus disease 2019 (COVID-19) pandemic, has led to more than 93 million cases and 2 million deaths globally as of January 17, 2021.[1] In response to the rapidly increasing burden, COVID-19 vaccine development has occurred at unprecedented speed, with several vaccines receiving authorizations or approvals worldwide in the first year after SARS-CoV-2 emergence.

The Moderna mRNA-1273 SARS-CoV-2 vaccine (Moderna COVID-19 vaccine, hereafter) is a lipid nanoparticle encapsulated messenger RNA (mRNA)-based vaccine encoding the prefusion-stabilized spike glycoprotein of SARS-CoV-2. The spike glycoprotein recognizes and binds to angiotensin-converting enzyme 2 receptors on host cells, facilitating viral entry.[7] The Moderna COVID-19 vaccine mimics natural infection by carrying genetic instructions to host cells to make the spike glycoprotein antigen, inducing T-cell and antibody responses. In early clinical trials, the Moderna COVID-19 vaccine demonstrated anti-SARS-CoV-2 immunogenicity and had an acceptable safety profile, with no trial-limiting severe adverse events.[8, 9]

A phase 3 randomized, observer-blinded, placebo-controlled trial of Moderna COVID-19 vaccine enrolled participants aged ≥18 years from July to October 2020 at 99 sites in the United States (U.S.). The trial enrolled approximately 30,000 participants who were randomized 1:1 to receive two intramuscular injections of Moderna COVID-19 vaccine or placebo 28 days apart. The incidence of symptomatic COVID-19 illness was 56.5 per 1000 person-years in the placebo group and 3.3 per 1000 person-years in the Moderna COVID-19 vaccine group.[6] Vaccine efficacy was 94.1% and was generally consistent across age, sex, race/ethnicity, and risk status.

Moderna is currently conducting a Phase 2/3 study to evaluate the safety and efficacy of its COVID-19 vaccine in healthy adolescents aged 12 to 17 years. Initial analyses of 3,235 participants showed vaccine efficacy of 96% with no serious safety concerns. [10]

Multiple SARS-CoV-2 variants of concern (VOCs) and variants of interest (VOIs) have also been identified over the course of the pandemic, some of which have been associated with greater disease severity and/or transmissibility. [2-4] In the U.S., VOCs include B.1.1.7 (Alpha),

B.1.351 (Beta), B.1.617.2 (Delta), and P.1 (Gamma), and VOIs include B.1.427 and B.1.429 (Epsilon), B.1.525 (Eta), B.1.526 (Iota), B.1.617.1 (Kappa), and B.1.617.3 (as of 07/27/2021). [11]

Following release of the phase 3 results in adults, the Moderna COVID-19 vaccine received Emergency Use Authorization (EUA) in the U.S. on December 18, 2020. COVID-19 vaccination was implemented in a phased approach, per Advisory Committee on Immunization Practices (ACIP) recommendations and state guidelines. In this protocol, we detail an observational cohort study to evaluate real-world vaccine effectiveness and durability of Moderna COVID-19 vaccine among a diverse population at Kaiser Permanente Southern California (KPSC).

### 3 Research Objectives

We propose to evaluate the vaccine effectiveness (VE) of Moderna COVID-19 vaccine in preventing COVID-19 diagnosis (symptomatic and asymptomatic) and severe COVID-19 disease (hospitalizations and mortality) at KPSC with the primary objectives below. Secondary objectives will be assessed but may be underpowered.

#### 3.1 Primary Objectives

1. To evaluate the effectiveness of 2 doses of Moderna COVID-19 vaccine in preventing COVID-19 diagnosis
2. To evaluate the effectiveness of 2 doses of Moderna COVID-19 vaccine in preventing severe COVID-19 disease

#### 3.2 Secondary Objectives

1. To evaluate the effectiveness of 2 doses of Moderna COVID-19 vaccine in preventing COVID-19 diagnosis by age and by sex
2. To evaluate the effectiveness of 2 doses of Moderna COVID-19 vaccine in preventing COVID-19 diagnosis by race/ethnicity groups
3. To evaluate the effectiveness of 2 doses of Moderna COVID-19 vaccine in preventing COVID-19 diagnosis in individuals with chronic diseases (e.g., chronic kidney disease, lung disease including chronic obstructive pulmonary disease [COPD] and asthma, diabetes)
4. To evaluate the effectiveness of 2 doses of Moderna COVID-19 vaccine in preventing COVID-19 diagnosis in individuals who are immunocompromised (e.g., HIV, cancer, transplant, immunosuppressive medications)
5. To evaluate the effectiveness of 2 doses of Moderna COVID-19 vaccine in preventing COVID-19 diagnosis in individuals with autoimmune conditions (e.g., rheumatoid arthritis, inflammatory bowel disease, psoriasis, psoriatic arthritis, multiple sclerosis, systemic lupus erythematosus)
6. To evaluate the effectiveness of 2 doses of Moderna COVID-19 vaccine in preventing COVID-19 diagnosis in frail individuals
7. To evaluate the effectiveness of 2 doses of Moderna COVID-19 vaccine administered during pregnancy in preventing COVID-19 diagnosis
8. To evaluate the effectiveness of 2 doses of Moderna COVID-19 vaccine in preventing COVID-19 diagnosis among individuals with a history of COVID-19 diagnosis
9. To evaluate the effectiveness of 2 doses of Moderna COVID-19 vaccine in preventing COVID-19 diagnosis when given concomitantly with another vaccine
10. To evaluate the effectiveness of 2 doses of Moderna COVID-19 vaccine in preventing asymptomatic COVID-19
11. To evaluate the effectiveness of 2 doses of Moderna COVID-19 vaccine in preventing symptomatic COVID-19
12. To evaluate the durability of 2 doses of Moderna COVID-19 vaccine in preventing COVID-19 diagnosis
13. To evaluate the durability of 2 doses of Moderna COVID-19 vaccine in preventing severe COVID-19 disease
14. To evaluate the effectiveness of 1 dose of Moderna COVID-19 vaccine in preventing COVID-19 diagnosis
15. To evaluate the effectiveness of 1 dose of Moderna COVID-19 vaccine in preventing severe COVID-19 disease
16. Assess the VE of two doses of Moderna COVID-19 vaccine against SARS-CoV-2 variants (using both test-negative and cohort designs)
17. Assess the VE of one dose of Moderna COVID-19 vaccine against SARS-CoV-2 variants (test-negative design only)

### 4 Study Setting

KPSC is one of the largest not-for-profit health plans and integrated health care systems in the U.S., providing an ideal environment for population-based research. KPSC’s population includes more than 4.5 million members, of which 702,136 adults are aged ≥65 years and 341,464 are between 12 and 17 years old (Table 1). The diverse demographic makeup, including 260 different ethnicities and more than 150 different languages, closely mirrors the Southern California population. Compared to the racial/ethnic distribution of the U.S. population, KPSC membership is composed of twice as many individuals of Asian/Pacific Island descent and three times as many Hispanic individuals.[12]

KPSC facilities include hospitals and medical offices, all linked by an information infrastructure that supports both clinical practice and business needs. Health information from this infrastructure can be leveraged for research purposes. More than 90 percent of members remain in the health plan after one year; more than three-quarters remain after three years. The large, diverse, and stable population permits the rapid accrual of a representative sample size and offers the ability to evaluate long-term implications of immunization.

Kaiser Permanente HealthConnect® is the largest and most advanced civilian electronic health record system available in the U.S. In addition to supporting patient care, this robust system facilitates research, providing access to electronic health records (EHR) for the research team. The medical record number serves as a unique identifier linking all medical encounters for each member. Care received in the outpatient, inpatient, and emergency settings is documented in the EHR and captured in research databases. Care received outside the KPSC system is captured through claims. Details of care are available at the fingertips of researchers in near real time.

Our EHR include a variety of data that can be used for research:

**Membership**: Includes demographic information such as sex, date of birth, and race/ethnicity.

**Diagnosis**: Includes International Classification of Diseases, 10th revision (ICD-10) codes.

**Procedure**: Includes ICD-10, Current Procedural Terminology (CPT), and Systematized Nomenclature of Medicine (SNOMED) codes.

**Immunization**: Includes vaccine name, date of vaccination, route of administration, facility where vaccine was administered, dose, manufacturer, and lot number.

**Laboratory**: Includes laboratory orders and results.

**Pharmacy**: Includes National Drug Codes (NDC) and Generic Product Identifier (GPI) codes.

More than 95 percent of members have a drug benefit with minimal copayments.

**Mortality**: Includes deaths from hospital and membership databases, as well as from state and national death files.

**Birth**: Includes pregnancy related information such as gestational age, birth weight, and Apgar scores.

**Clinical notes**: Allows for natural language processing of clinical notes to identify outcomes not easily identified through structured data.

##### Table 1. Demographic characteristics of Kaiser Permanente Southern California members on January 1, 2021

|  | **Number of members** |
| --- | --- |
| Total population | 4,573,594 |
| **Sex** |  |
| Male | 2,360,957 |
| Female | 2,212,637 |
| **Age (years)** |  |
| <6 | 278,559 |
| 6 to <12 | 308,704 |
| 12 to <18 | 341,464 |
| 18 to <65 | 2,942,731 |
| 65 to <75 | 432,045 |
| ≥75 | 270,091 |
| **Race** |  |
| White | 2,470,912 |
| Black or African American | 359,270 |
| American Indian & Alaska Native | 21,137 |
| Asian | 487,947 |
| Native Hawaiian and Other Pacific Islander | 37,107 |
| Other race | 1,172,221 |
| Two or more races | 25,000 |
| **Ethnicity** |  |
| Hispanic or Latinx (of any race) | 1,829,983 |

#### 4.1 COVID-19 Vaccination at KPSC

Immunizations are an important part of KPSC’s overall focus on preventive care. The organization is one of the top-rated health maintenance organizations for meeting national standards of care, which include measures of childhood and adult immunization. Recommended vaccines are provided at no cost to KPSC members. KPSC thus provides an excellent real- world setting in which to understand the effectiveness of vaccines used in the course of routine clinical care.

According to California’s COVID-19 Vaccination Plan from October 2020, the state would allocate vaccines directly to large multi-jurisdictional entities, such as health providers and systems with locations in multiple counties. A multi-county entity (MCE) is a health system that has facilities in more than two California counties to centrally support local implementation in all of its locations, set policy for all of its facilities, order and store vaccine, has a centralized

pharmacy, and has a demonstrated track record in immunizing their staff. KPSC is one of the largest California MCEs.

In December 2020, KPSC began to administer Moderna COVID-19 vaccine to eligible individuals aged ≥18 years and Pfizer-BioNTech COVID-19 vaccine to eligible individuals aged

≥16 years. In March 2021, KPSC began to administer Janssen COVID-19 vaccine to eligible individuals aged ≥18 years. KPSC is able to differentiate COVID-19 vaccine products in the EHR.

Once Moderna COVID-19 vaccine is authorized and recommended for individuals 12 to 17 years of age, KPSC will administer Moderna COVID-19 vaccine to this population and they will be included in this study.

To ensure equitable distribution, California was allocating COVID-19 vaccines as they became available. KPSC followed the state’s COVID-19 vaccine prioritization. Individuals were prioritized for vaccination as follows:

- Phase 1A (started in December 2020)
  - Healthcare workers
  - Long-term care residents
- Phase 1B (started in January 2021)
  - Individuals aged ≥65 years
  - Sector populations:
    - Education and childcare
    - Emergency services
    - Food and agriculture
- Individuals aged 16-64 years (started in March 2021)
  - At the very highest risk for morbidity and mortality from COVID-19 as a direct result of one or more of the severe health conditions
- Individuals aged 16 years and older (started in April 2021)
- Individuals aged 12 years and older (started in May 2021)

KPSC is an approved COVID-19 vaccine provider and is receiving Moderna COVID-19 vaccine from the state of California. KPSC also received Moderna COVID-19 vaccine directly from Moderna as part of this real-world effectiveness study.

#### 4.2 COVID-19 at KPSC

At KPSC, diagnostic testing for SARS-CoV-2 is offered free of charge with an order from a KPSC physician, which can be requested by email, in person or through a virtual visit.

Prioritization for testing has evolved during the pandemic, with an initial emphasis on individuals with symptoms (particularly high-risk groups) and prior to hospital admissions or certain outpatient procedures, with gradual expansion in 2020 to members without COVID-19 associated symptoms.

Testing in 2020 was primarily conducted by RT-PCR of nasopharyngeal/oropharyngeal swabs using the Roche cobas® SARS-CoV-2 assay on the Roche cobas® 6800 and 8800 analyzers or nasal/oropharyngeal swabs using the Aptima® SARS-CoV-2 assay on the Hologic Panther® analyzers. A small number of Abbott ID NOW™ COVID-19 rapid tests were conducted in limited settings (e.g., obstetrics, pulmonary medicine, and infectious disease departments); these were phased out by end of 2020. In November 2020, KPSC opened a new regional COVID-19

laboratory with Thermo Fisher Scientific Amplitude Solution instruments and also added saliva testing for asymptomatic individuals, increasing testing capacity to approximately 52,000 tests per day. In January 2021, voluntary weekly saliva testing was introduced for physicians and employees working in patient-facing areas.

As of July 2021, the TaqPath™ COVID-19 High-Throughput Combo Kit on the Thermo Fisher Scientific Amplitude Solution is used for the majority of SARS-CoV-2 molecular tests. A small proportion of tests are conducted using the Roche cobas® SARS-CoV-2 assay on the Roche cobas® 8800 System or the Roche cobas® SARS-CoV-2 & Influenza A/B assay on the Roche cobas® Liat® System; these tests can be ordered in limited circumstances when a provider requires an expedited result. The Aptima® SARS-CoV-2 assay on the Hologic Panther® System was phased out in early 2021.

##### Table 2. SARS-CoV-2 molecular tests and diagnoses among members of Kaiser Permanente Southern California during 01/01/2020-12/31/2020

|  | **Age (years)** | | | **Total** |
| --- | --- | --- | --- | --- |
|  | **12-17** | **18-64** | **≥65** |  |
| Number of patients tested for SARS-CoV-2 with RT-PCR test | 48,290 | 825,670 | 184,197 | 1,058,157 |
| Number of patients with a positive SARS-CoV-2 RT-PCR test | 13,788 | 200,403 | 24,452 | 238,643 |
| Number of patients with a COVID-19 diagnosis^1^ | 15,807 | 247,628 | 31,960 | 295,395 |
| Number of patients admitted to hospital with a COVID-19 diagnosis^1^ | 87 | 10,871 | 7,980 | 18,938 |
| Number of patients admitted to ICU with a COVID-19 diagnosis^1^ | 13 | 2,928 | 2,227 | 5,168 |
| Number of deaths within 31 days after the first COVID-19 diagnosis^1^ | 1 | 715 | 2,038 | 2,754 |

^1^A COVID-19 diagnosis includes those with a positive SARS-CoV-2 RT-PCR test result or a COVID-19 diagnosis code only.

As of December 31, 2020, over 1 million SARS-CoV-2 RT-PCR tests had been performed at KPSC, of which 17.4% were among individuals aged ≥65 years and 4.6% were among adolescents aged 12-17 years (Table 2). There were 238,643 members testing positive (13,788 members aged 12-17 years and 24,452 members aged ≥65 years). Additional members were diagnosed with COVID-19 based on positive test results outside of KPSC or clinical presentation and contact history, for a total of 56,752 members with a COVID-19 clinical diagnosis only. Approximately 25.0% (7,980/31,960) of COVID-19 patients aged ≥65 years were hospitalized, compared to only 4.4% (10,871/247,628) who were hospitalized among patients aged 18-64 years and 0.6% (87/15,807) who were hospitalized among patients aged 12-17 years. The proportion of COVID-19 patients being admitted to the ICU was 7.0% in patients aged ≥65 years versus 1.2% in patients aged 18-64 years. The proportion of COVID-19 patients who died within 31 days after a COVID-19 diagnosis or a positive test was 6.4% in patients aged ≥65 years versus 0.29% in patients aged 18-64 years.

As of July 2021, asymptomatic individuals who receive a COVID-19 test at KPSC may include individuals without COVID-19 symptoms who require or request testing. Both vaccinated and unvaccinated individuals are required to be tested for SARS-CoV-2 prior to KPSC procedures or admission. In addition, asymptomatic individuals can request testing (regardless of vaccination status) for any of the following reasons: travel, exposure to or close contact with a COVID-19-

positive individual, residents or employees of congregate living facilities, employment (e.g. healthcare workers, first responders, essential workers, or any others who require testing for workplace), school or daycare, or any other reason. In addition, asymptomatic individuals who are physicians or other employees of KPSC can receive voluntary weekly saliva testing. Testing for asymptomatic individuals most commonly uses saliva samples rather than nasal/oropharyngeal swabs.

#### 4.3 SARS-CoV-2 Whole Genome Sequencing

KPSC began saving positive molecular SARS-CoV-2 test specimens in March 2021 to conduct whole genome sequencing (WGS). As part of this effort, all positive SARS-CoV-2 molecular test specimens from March 2021 to February 2022 will be sent to an external laboratory for sequencing and variant identification.

### 5 Study Design Overview

We will conduct an observational cohort study to evaluate the VE of Moderna COVID-19 vaccine in preventing COVID-19 diagnosis and severe COVID-19. Vaccine exposure, COVID- 19 outcomes, and covariates will be identified from EHR, as described in Section 7 below. All individuals who receive Moderna COVID-19 vaccine from December 2020 to December 2021 and meet eligibility criteria specified in Section 6.1 below will be included in the study (“vaccinated”, hereafter). The unvaccinated comparator cohort will comprise individuals who have not received Moderna COVID-19 vaccine or any other COVID-19 vaccine as of the index date of their matched vaccinated individual (see Section 6.2 for details on index date) and who meet eligibility criteria specified in Section 6.1 below. Unvaccinated individuals who subsequently receive Moderna COVID-19 vaccine may contribute person-time to both unvaccinated and vaccinated cohorts. Individuals will be followed up through EHR for occurrence of COVID-19 outcomes until the end of the study period (December 31, 2023 for final analysis), or censoring events (termination of KPSC membership allowing for a 31-day gap, death, receipt of a COVID-19 vaccine).

Six interim analyses and a final analysis will be conducted. For the first and second interim analyses, unvaccinated individuals will be randomly selected and n:1 matched to vaccinated individuals by age, sex, and race/ethnicity (if sample size allows), and will be assigned an index date based on the vaccination date of their matched vaccinated individual. For subsequent interim analyses and the final analysis, the number of eligible unvaccinated individuals in the KPSC population may be insufficient for individual matching; if so, frequency matching will be employed. In all analyses, efforts will be dedicated to identifying and adjusting for potential confounders. Cox proportional hazards regression will be used to estimate unadjusted and adjusted HRs. VE (%) will be estimated as (1 – adjusted HR) × 100. Propensity score analyses with inverse probability of treatment weighting (IPTW) will be considered for final analyses.

Analyses will also be conducted for secondary objectives to estimate VE of 2 doses of Moderna COVID-19 vaccine in sub-populations of interest, to estimate VE of 2 doses of Moderna COVID- 19 vaccine when received concomitantly with other vaccines, to estimate VE of 2 doses of Moderna COVID-19 vaccine in preventing asymptomatic COVID-19 and symptomatic COVID- 19, to estimate durability of 2 doses of Moderna COVID-19 vaccine, and to estimate VE of 1 dose of Moderna COVID-19 vaccine.

We will also conduct analyses to assess VE against specific SARS-CoV-2 variants (Secondary Objectives 16-17). The test-negative and cohort study designs and analytic plan for these analyses are detailed in Section 9 below.

### 6 Study Population

6.1 Eligibility Criteria

Individuals will be eligible for inclusion in the study if they meet the criteria below. Index dates are detailed in Section 6.2 below.

###### Inclusion criteria

- Aged ≥12 years at index date
- KPSC member for ≥12 months prior to index date through 14 days after the index date (allowing a 31-day gap)

Membership is necessary to follow-up individuals for COVID-19 outcomes in the EHR. In addition, prior membership allows comorbidities and other covariates to be identified and considered in analyses as potential confounders.

###### Exclusion criteria

- Receipt of a COVID-19 vaccine other than Moderna COVID-19 vaccine prior to or on the index date
- Receipt of 2 doses of Moderna COVID-19 vaccine <24 days apart [13] for 2-dose exposed cohort
- Receipt of any COVID-19 vaccine <14 days after the index date
- No health care utilization and no vaccination from the 2 years prior to the index date through the index date
- Occurrence of a COVID-19 outcome <14 days after the index date
  6.2 Index Dates

All individuals meeting criteria in Section 6.1 will be included in analyses. Index dates will be assigned at each analysis. For vaccinated individuals, the index date for Primary Objectives 1-2 and Secondary Objectives 1-13 will be the date of receipt of the second dose of Moderna COVID-19 vaccine. For Secondary Objectives 14-15 among individuals who receive only 1 dose of Moderna COVID-19 vaccine, the index date for vaccinated individuals will be the date of receipt of the first dose of Moderna COVID-19 vaccine.

The index date for the comparison group will depend on the analysis. The comparison group will comprise a similar population meeting criteria in Section 6.1 who are unvaccinated at the index date. For the first and second interim analyses (if sample size allows), unvaccinated individuals will be randomly selected and n:1 matched to the vaccinated individuals by age (12-17 years, 18-44 years, 45-64 years, 65-74 years, and ≥75 years), sex, and race/ethnicity (Non-Hispanic White, Non-Hispanic Black, Hispanic, Non-Hispanic Asian, and Other/Unknown). The matching ratio will depend on the sample size of the unvaccinated population, between 1:1 to 5:1. The index date for the unvaccinated match will be the same as his/her matched vaccinated counterpart.

For the subsequent interim analyses and final analysis, we expect that there will be insufficient unvaccinated individuals for 1:1 matching, as COVID-19 vaccination is expanded to the general

population. If this is the case, unvaccinated individuals will be randomly selected and matched to the vaccinated individuals by the distribution (frequency) of age, sex, and race/ethnicity of vaccinated individuals. Since frequency matching is not conducted on the individual level, an index date will be assigned to unvaccinated individuals based on the distribution of calendar time of vaccinated individuals on the vaccination (index) date in each age, sex, and race/ethnicity stratum. For example, if 5% of vaccinated individuals are vaccinated during the week of February 1, 2021, 5% of unvaccinated individuals will be assigned an index date of February 1, 2021. Hence, the distribution of index dates between the vaccinated group and the unvaccinated group will be matched. For each interim analysis during the accrual period (December 2020 to December 2021), the comparison group will be assessed and assigned with index dates independently based on all available data.

### 7 Measures

#### 7.1 Exposure Definition

The primary exposure for this study (Primary Objectives 1-2, Secondary Objectives 1-13) will be receipt of 2 doses of Moderna COVID-19 vaccine received ≥24 days apart (allowing a 4-day grace period prior to the recommended interval of 28 days) during the accrual period. The exposure for Secondary Objectives 14-15 will be 1 dose of Moderna COVID-19 vaccine among individuals who received only 1 dose during the accrual period, and the exposure for Secondary Objective 9 will be receipt of 2 doses of Moderna COVID-19 vaccine during the accrual period, with either dose given concomitantly with another vaccine.

- CVX code: 207
- KPSC Immunization ID and description:
  - 124285 COVID-19, mRNA, LNP-S, PF (Moderna)
  - 124287 COVID-19 vaccine, Moderna, external administration
- Manufacturer: Moderna, Inc
- Product name: Moderna COVID-19 Vaccine
- NDC 10/NDC 11 Labeler Product ID (vial): 80777-273-10 or 80777-0273-10

#### 7.2 Outcome Definitions

The primary outcomes for this study are:

1. COVID-19 diagnosis will be defined as a SARS-CoV-2 positive molecular test or a COVID-19 diagnosis code (Primary Objective 1, Secondary Objectives 1-12, 14)
2. Severe COVID-19 disease includes COVID-19 hospitalization (hospitalization with a SARS-CoV-2 positive test or a COVID-19 diagnosis, or a hospitalization occurring ≤7 days after a SARS-CoV-2 positive test, with chart review to confirm severe COVID-19 symptoms) and COVID-19 mortality (death during COVID-19 hospitalization) (Primary Objective 2, Secondary Objectives 13, 15)

We will ascertain the first occurrence of COVID-19 diagnosis or severe COVID-19 disease ≥14 days after the index date.

Incident COVID-19 cases (identified through diagnosis code or positive molecular test) will be separated into symptomatic and asymptomatic COVID-19 cases. COVID-19 symptoms will be identified using a natural language processing (NLP) algorithm (currently being submitted for publication). The NLP algorithm will be applied to clinical notes before and after the incident COVID-19 diagnosis date to search for COVID-19 related symptoms. COVID-19 cases with no symptoms identified will be considered asymptomatic cases.

Positive SARS-CoV-2 tests conducted for surveillance of asymptomatic individuals will also be used as a preliminary approach to identify asymptomatic COVID-19 cases until the NLP algorithm is finalized. The use of testing data in asymptomatic individuals will facilitate assessment of VE against asymptomatic COVID-19 for the first interim analysis, whereas the

more comprehensive NLP approach will require more time to apply and will be used for subsequent analyses.

#### 7.3 Other Variable Definitions

Other variables (Table 3) will be identified from EHR and considered in analyses when feasible and appropriate as covariates or stratification variables.

##### Table 3. List of possible covariates

|  | **Categories** |
| --- | --- |
| **Variables to be assessed at index date (Day 0)** | |
| Age at index date, years | 12-17, 18-44, 45-64, 65-74, 75+ |
| Sex | Female, Male |
| Race/Ethnicity | Non-Hispanic White, Non-Hispanic Black, Non-Hispanic Asian, Hispanic, Other/Unknown |
| Month of index date | By calendar month |
| **Socioeconomic status** |  |
| Medicaid | Yes, No |
| Neighborhood median household income | To be determined based on variable distribution |
| Medical center area | MC1, MC2, MC3, … |
| Pregnancy status | Yes (1st trimester, 2nd trimester, 3rd trimester), No |
| **Variables to be assessed in the two years prior to index date (-730 to -1 day, inclusive)** | |
| Any smoking behavior | Yes, No, Unknown |
| Most recent Body Mass Index (BMI) measurement | <18.5, 18.5 - <25, 25 - <30, ≥30 - <35, 35 -  <40, 40 - <45, ≥45, Unknown |
| **Variables to be assessed in the year prior to index date (-365 to -1 day, inclusive)** | |
| Charlson comorbidity score [14]* | 0, 1, 2+ |
| **Autoimmune conditions** | Yes, No |
| Rheumatoid arthritis | Yes, No |
| Inflammatory bowel disease | Yes, No |
| Psoriasis | Yes, No |
| Psoriatic arthritis | Yes, No |
| Multiple sclerosis | Yes, No |
| Systemic lupus erythematosus | Yes, No |
| **Health care utilization** |  |
| Number of virtual and outpatient encounters | 0, 1-4, 5-10, 11+ |
| Number of emergency encounters | 0, 1, 2+ |
| Number of inpatient encounters | 0, 1, 2+ |
| Preventive care - with other vaccinations, screenings, and well-visits from all settings | Yes, No |

|  | **Categories** |
| --- | --- |
| **Chronic diseases** |  |
| Kidney disease | Yes, No |
| Heart disease | Yes, No |
| Lung disease | Yes, No |
| Liver disease | Yes, No |
| Diabetes | Yes, No |
| **Other variables** | |
| KPSC physician/employee status at index date | Yes, No |
| Frailty index in year prior to index date (-365 to -1 day, inclusive), using method by Kim et al. [15] | To be determined based on quartiles |
| History of COVID-19 diagnosis (from March 1, 2020 to index date, inclusive) | Yes, No |
| History of SARS-CoV-2 molecular test performed (from March 1, 2020 to index date, inclusive), regardless of result | Yes, No |
| Concomitant vaccine | Yes, No |
| **Immunocompromised status** | Yes, No |
| HIV/AIDS any time prior to index date | Yes, No |
| Leukemia, lymphoma, congenital immunodeficiencies, asplenia/hyposplenia any time prior to index date | Yes, No |
| Organ transplant any time prior to index date | Yes, No |
| Immunosuppressant medications at index date | Yes, No |

* Charlson comorbidities include myocardial infarction, congestive heart failure, peripheral vascular disease, cerebrovascular disease, connective tissue disease, dementia, chronic pulmonary disease, connective tissue disease, peptic ulcer disease, diabetes mellitus, paraplegia and hemiplegia, renal disease, liver disease, cancer, metastatic solid tumor, human immunodeficiency virus (HIV), and acquired immunodeficiency syndrome (AIDS)

The covariates included in adjusted analyses will be determined by scientific relevance, association with exposure, and data availability. Specifically, we will select covariates by following these steps:

1. The distribution of covariates will be reviewed. Some categories of a categorical variable with small sample sizes may be redefined or combined into one category.
2. The association of baseline covariates with exposure will be assessed. We will use standardized difference to assess the balance of covariates between exposed and unexposed cohorts. Unlike p-values, for which magnitude is highly related to sample size, standardized difference is a unified approach to quantifying the magnitude of difference between groups regardless of sample size, where an absolute value less than 0.1 is considered a negligible difference. Potential confounders will be determined by absolute standardized difference (ASD) >0.1.
3. All potential confounders from Step 2 will be included in the analyses. Steps 1 and 2 will be repeated for the 2-dose cohort and the 1-dose cohort. Matching variables (age, sex, race/ethnicity and index date), which are considered important risk factors, will be kept in the

adjusted model for possible imbalance on matching variables due to loss to follow-up or subgroup analysis (e.g., by baseline comorbidities).

### 8 Statistical Analysis for Study Objectives 1-15

#### 8.1 Analysis Plan

For Primary Objectives 1-2, interim analyses will be conducted every 3 months in the first year, beginning in July 2021 (Figure 1). The first interim analysis will include all vaccinated individuals who are accrued in the first ~3 months of the study (December 2020 to March 2021) and followed for at least 3 months after completion of the second dose. The second and third interim analyses will be performed in October 2021 and January 2022 with cumulative accrual from December 2020 to June 2021 and December 2020 to September 2021, respectively. Interim analyses will be performed every 6 months in the second and the third year. Matching of the unvaccinated comparator cohort to the vaccinated cohort will be conducted separately at each of the first 4 interim analyses, such that each analysis will have a different unvaccinated cohort. However, the unvaccinated cohort will remain fixed beginning with the 4th interim analysis through the final analysis (Primary Objectives 1-2 and Secondary Objectives 1-15), which will include all vaccinated individuals accrued in 2021, with follow-up through December 2023. No data lags are applied for the first 3 interim analyses, but a 3-month data lag is applied starting with the 4th interim analysis. The removal of the data lag for the first 3 interim analyses will permit more rapid generation of results. Without the 3-month data lag, some events reported through claims may be missed, leading to an underestimation of incidence rates. However, we expect the proportion of claims to be non-differential in the vaccinated and unvaccinated cohorts.

##### Figure 1. Analysis Plan for Study Objectives 1-15

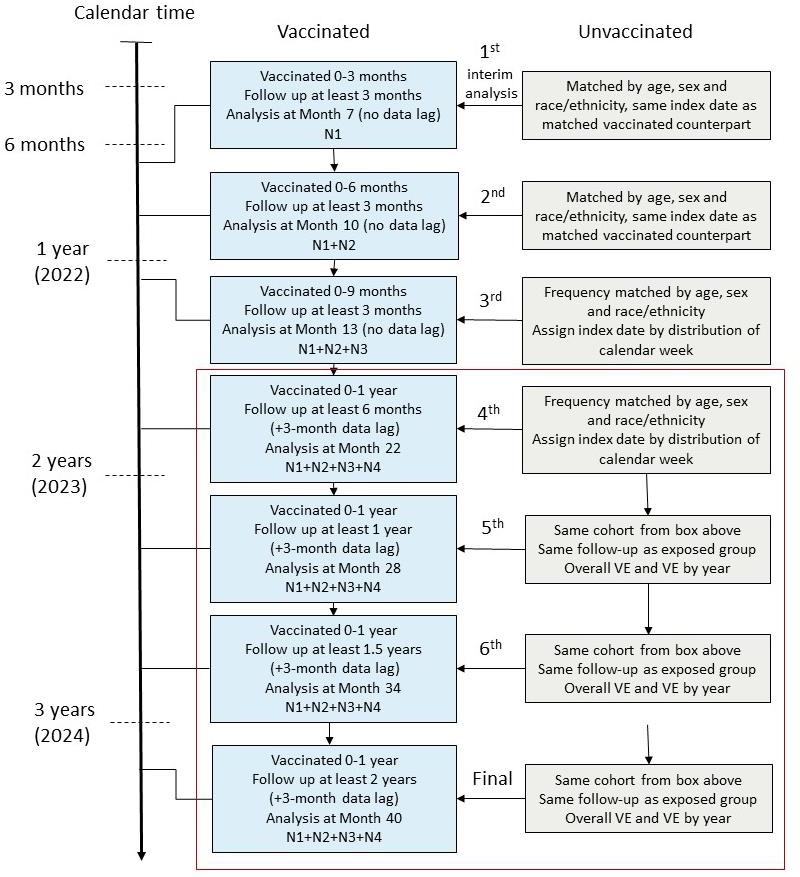

#### 8.2 Descriptive Analysis

Descriptive attributes of vaccinated and unvaccinated cohorts will be presented as absolute numbers and percentages. We will use a χ2 test to test for significant differences in the distribution of the categorical covariates among individuals between each of the vaccinated and unvaccinated cohorts at cohort entry. Continuous variables will be presented as the mean with standard deviation and/or median with interquartile ranges, with p-values for the two-sample t- test or Wilcoxon rank-sum test, as appropriate. Absolute standardized differences will be calculated to assess the balance of covariates.

Overall incidence rates of COVID-19 diagnosis and of severe COVID-19 for the vaccinated and unvaccinated cohorts will be calculated by dividing the number of incident events by the total number of person-years.

#### 8.3 Primary Analyses

The primary analyses addressing Primary Objectives 1-2 will be conducted at interim and final analyses (Figure 1).

Unadjusted hazard ratios (HR) and adjusted HR and confidence intervals (CIs) comparing COVID-19 diagnosis or severe COVID-19 disease in vaccinated and unvaccinated individuals will be estimated by Cox proportional hazards regression models without and with adjustment for potential confounders described in Section 7.3. Unadjusted VE (%) will be calculated as (1- unadjusted HR) x 100. Adjusted VE (%) will be calculated as (1 – adjusted HR) × 100. For final analysis of primary objectives, we may also conduct propensity score analyses with inverse probability of treatment weighting (IPTW) to balance covariates across exposure groups.

For each interim analysis and final analysis, the test significance level will be adjusted using an alpha spending approach without a stopping rule (similar to the Bonferroni correction, p=0.007=0.05/7). This is a conservative approach to keep the overall Type I error below 0.05. As such, we will be calculating 99.3% CIs for each interim and final analysis. We will provide 95% CIs for each analysis as well.

#### 8.4 Secondary Analyses

Secondary analyses will be conducted for the 4th interim and the final analysis. Select secondary analyses will also be conducted for the 1st interim analysis (VE by age, sex, race/ethnicity, history of COVID-19, and asymptomatic COVID-19 [based on individuals with a positive result from a surveillance/asymptomatic test]) and for the 2nd interim analysis (VE in pregnant women).

For each secondary analysis, we will examine the distribution of index dates in vaccinated individuals; we will assign index dates by stratified variables so that the frequency distribution of their index dates matches that of vaccinated individuals.

***Secondary Objective 1:*** For Secondary Objective 1, analyses will be stratified by sex and age category (12-17 years, 18-44 years, 45-64 years, 65-74 years, and 75+ years) at index date.

For each stratum, we will calculate COVID-19 diagnosis incidence, unadjusted and adjusted HRs and VEs, as described for the primary analysis above.

***Secondary Objective 2:*** For Secondary Objective 2, analyses will be stratified by race/ethnicity (Non-Hispanic White, Non-Hispanic Black, Hispanic, and Non-Hispanic Asian). For each stratum, we will calculate COVID-19 diagnosis incidence, unadjusted and adjusted HRs and VEs, as described for the primary analysis above.

***Secondary Objective 3:*** For Secondary Objective 3, analyses will include individuals with chronic diseases (e.g., chronic kidney disease, lung disease including COPD and asthma, diabetes, identified by ICD-10 codes). For each disease, we will calculate COVID-19 diagnosis incidence, unadjusted and adjusted HRs and VEs, as described for the primary analysis above.

***Secondary Objective 4:*** For Secondary Objective 4, analyses will include individuals who are immunocompromised (e.g., HIV, cancer, transplant, immunosuppressive medications identified by ICD-10 codes, registries, and pharmacy data). We will calculate COVID-19 diagnosis incidence, unadjusted and adjusted HRs and VEs, as described for the primary analysis above.

***Secondary Objective 5:*** For Secondary Objective 5, analyses will include individuals with autoimmune conditions (e.g., rheumatoid arthritis, inflammatory bowel disease, psoriasis, psoriatic arthritis, multiple sclerosis, systemic lupus erythematosus). We will calculate COVID- 19 diagnosis incidence, unadjusted and adjusted HRs and VEs, as described for the primary analysis above.

***Secondary Objective 6:*** For Secondary Objective 6, analyses will include individuals who are frail, identified using the method by Kim et al.[15] We will calculate COVID-19 diagnosis incidence, unadjusted and adjusted HRs and VEs, as described for the primary analysis above.

***Secondary Objective 7:*** For Secondary Objective 7, analyses will include pregnant women. We will calculate COVID-19 diagnosis incidence, unadjusted and adjusted HRs and VEs, as described for the primary analysis above.

***Secondary Objective 8:*** For Secondary Objective 8, analyses will include individuals with a history of COVID-19 diagnosis. Incident COVID-19 diagnosis (a COVID-19 reinfection, >90 days after the most recent prior COVID-19 diagnosis code or SARS-CoV-2 positive molecular test) during follow-up will be assessed using two definitions. The first definition will require a COVID- 19 diagnosis code with chart-confirmed symptoms or a SARS-CoV-2 positive molecular test.

The second, more specific definition will require a COVID-19 diagnosis code with chart- confirmed symptoms, a SARS-CoV-2 positive molecular test with chart-confirmed symptoms, or a SARS-CoV-2 positive molecular test with an intervening SARS-CoV-2 negative molecular test. We will calculate COVID-19 diagnosis incidence, unadjusted and adjusted HRs and VEs, as described for the primary analysis above.

***Secondary Objective 9:*** CDC initially recommended that COVID-19 vaccines be routinely administered alone, with a minimum interval of 14 days before or after administration with any other vaccine. In May 2021, CDC updated their recommendations to indicate that COVID-19 vaccines and other vaccines may be administered without regard to timing; this includes administration of COVID-19 vaccines and other vaccines on the same day as well as within 14 days of each other. For Secondary Objective 9, we will compare individuals who received either the first or second dose of Moderna COVID-19 vaccine concomitantly with another vaccine (i.e., on the same day) with unvaccinated individuals. We will calculate COVID-19 diagnosis incidence and unadjusted and adjusted HRs and VEs, as described for the primary analysis above.

***Secondary Objective 10:*** For Secondary Objective 10, we will calculate VE of 2 doses of Moderna COVID-19 vaccine against asymptomatic COVID-19. We will calculate the incidence of asymptomatic COVID-19 and estimate unadjusted and adjusted HRs and VEs, as described for the primary analysis above.

***Secondary Objective 11:*** For Secondary Objective 11, we will calculate VE of 2 doses of Moderna COVID-19 vaccine against symptomatic COVID-19. We will calculate the incidence of symptomatic COVID-19 and estimate unadjusted and adjusted HRs and VEs, as described for the primary analysis above.

***Secondary Objective 12:*** For Secondary Objective 12, we will estimate the VE against COVID- 19 diagnosis by year. We will calculate COVID-19 diagnosis incidence by follow-up year by dividing the number of incident events occurring within that year by the number of person-years of follow-up for individuals at risk in that year. We will use time-varying Cox regression models to estimate unadjusted and adjusted HRs and VEs, for follow-up Year 1, Year 2, and Year 3 after the index date.

***Secondary Objective 13:*** For Secondary Objective 13, we will estimate the VE against severe COVID-19 by year. We will calculate severe COVID-19 incidence, unadjusted and adjusted HRs and VEs, as described for the Secondary Objective 12 analysis above, for follow-up Year 1, Year 2, and Year 3 after the index date.

***Secondary Objective 14:*** For Secondary Objective 14, we will estimate the VE against COVID- 19 diagnosis among those who only received 1 dose of Moderna COVID-19 vaccine compared to unvaccinated individuals. We will calculate COVID-19 diagnosis incidence, unadjusted and adjusted HRs and VEs, as described for the primary analysis above.

***Secondary Objective 15:*** For Secondary Objective 15, we will estimate the VE against severe COVID-19 among those who only received 1 dose of Moderna COVID-19 vaccine compared to unvaccinated individuals. We will calculate severe COVID-19 incidence, unadjusted and adjusted HRs and VEs, as described for the primary analysis above.

### 9 Sample Size and Power Estimation

Power calculations depend on Moderna COVID-19 vaccine implementation, uptake, and the number of individuals meeting criteria for comparable vaccinated and unvaccinated cohorts.

We estimate that a total of 80,000 individuals will receive 2 doses of Moderna COVID-19 vaccine from December 2020 through March 2021, assuming 80% series completion (first interim analysis). We estimate an incidence of 30 COVID-19 diagnoses per 1,000 unvaccinated adult KPSC members during an average 4-month follow-up period (accounting for possible censoring). We expect to have >99.9% power to detect a VE of 70%, with various sample sizes in the unvaccinated group (Table 4), using a 2-sided test with alpha=0.007 (0.05 adjusted for 6 interim analyses and 1 final analysis). The calculation was performed using SAS software package (version 9.4) PROC POWER procedure.

##### Table 4. Power calculations for 1st interim analysis of Primary Objective 1 for various sample sizes and vaccine effectiveness estimates

| **Sample Size Ratio (Vaccinated vs**  **Unvaccinated)** | **Incidence rate in unvaccinated group**  **(cases/1000 persons)** | **VE (%)**  **(reduction rate)** | **Power** |
| --- | --- | --- | --- |
| 1 vs 0.5 | 30 | 90 | >.999 |
|  | 30 | 80 | >.999 |
|  | 30 | 70 | >.999 |
| 1 vs 1 | 30 | 90 | >.999 |
|  | 30 | 80 | >.999 |
|  | 30 | 70 | >.999 |
| 1 vs 5 | 30 | 90 | >.999 |
|  | 30 | 80 | >.999 |
|  | 30 | 70 | >.999 |

For the first interim analysis of Primary Objective 2, we estimate an incidence of 2 COVID-19 hospitalizations and 0.3 deaths per 1,000 unvaccinated adult KPSC members during an average 4-month follow-up period. We expect to have >99.9% power to detect a 70% VE against COVID-19 hospitalizations, with various sample sizes in the unvaccinated group (Table 5).

##### Table 5. Power calculations for 1st interim analysis of Primary Objective 2 for various sample sizes and vaccine effectiveness estimates

| **Sample Size Ratio**  **(Vaccinated vs Unvaccinated)** | **Incidence rate in**  **unvaccinated group (cases/1000 persons)** | **VE (%)**  **(reduction rate)** | **Power** |
| --- | --- | --- | --- |
| 1 vs 0.5 | 2 | 90 | >.999 |
|  | 2 | 80 | >.999 |
|  | 2 | 70 | >.999 |
| 1 vs 1 | 2 | 90 | >.999 |
|  | 2 | 80 | >.999 |
|  | 2 | 70 | >.999 |
| 1 vs 5 | 2 | 90 | >.999 |
|  | 2 | 80 | >.999 |
|  | 2 | 70 | >.999 |
| 1 vs 0.5 | 0.3 | 90 | 0.84 |
|  | 0.3 | 80 | 0.69 |
|  | 0.3 | 70 | 0.51 |
| 1 vs 1 | 0.3 | 90 | 0.93 |
|  | 0.3 | 80 | 0.81 |
|  | 0.3 | 70 | 0.62 |
| 1 vs 5 | 0.3 | 90 | 0.99 |
|  | 0.3 | 80 | 0.97 |
|  | 0.3 | 70 | 0.82 |

Based on the current Moderna COVID-19 vaccine implementation plan and anticipated uptake, we expect to accrue 500,000 to 1,000,000 individuals receiving 2 doses of Moderna COVID-19 vaccine for analyses of primary objectives by the end of 2021.

### 10 Statistical Analysis Plan for Vaccine Effectiveness Against SARS-CoV-2 Variants (Study Objectives 16-17)

As multiple SARS-CoV-2 variants of concern (VOCs) and variants of interest (VOIs) have been identified over the course of the pandemic, it is essential to identify the viral variant causing disease in this long-term effectiveness study. WGS of SARS-CoV-2 positive specimens from vaccinated and unvaccinated individuals will be performed to achieve the following secondary objectives:

16. Assess the VE of two doses of Moderna COVID-19 vaccine against SARS-CoV-2 variants (using both test-negative and cohort designs)
17. Assess the VE of one dose of Moderna COVID-19 vaccine against SARS-CoV-2 variants (test-negative design only)

To assess VE of 2 doses and 1 dose of Moderna COVID-19 vaccine against SARS-CoV-2 VOCs and VOIs (Secondary Objectives 16 and 17), we aim to use a test-negative design (TND). The TND includes all sequencing data and more easily facilitates both 2-dose and 1- dose VE analyses. In contrast, a cohort design includes all vaccinated individuals; however, the cohort analysis will only be conducted for the final analysis if the proportions of positive SARS- CoV-2 tests at KPSC and positive specimens sent for sequencing among COVID-19 cases (i.e., COVID-19 diagnosis outcome) are similar between vaccinated and unvaccinated groups; if the proportions are substantially different a cohort design will yield biased results. The threshold for testing individuals with symptoms suggestive of COVID-19 at KPSC does not differ by vaccination status. VOCs and VOIs will be selected for analyses based on prevalence in the KPSC population and scientific interest. Descriptive analyses will include the percentages of specimens from vaccinated and unvaccinated individuals sequenced with valid results, stratifying by cycle threshold (Ct) values, specimen type, and symptomatic/asymptomatic test types. We will also examine the proportions of vaccinated and unvaccinated individuals with each VOC and VOI overall, by specific characteristics (e.g., age, sex, race/ethnicity, immunocompromised status, other comorbidities, history of COVID-19), COVID-19 hospitalization/hospitalized death, and month of specimen collection.

#### 10.1 Test-negative design (TND)

##### Inclusion criteria

- Aged ≥18 years (aged ≥12 years once Moderna COVID-19 vaccine is authorized in this age group) as of date of specimen collection for the SARS-CoV-2 test (Day 0)
- KPSC member for ≥12 months prior to Day 0 (Day -365 to Day 0, inclusive, and allowing a 31-day gap)
- Tested positive for SARS-CoV-2 by PCR at KPSC with specimen collected on or after 3/1/2021 and sent for WGS, without a history of a COVID-19 diagnosis code or SARS- CoV-2 positive molecular test in the 90 days prior to the test; or tested negative for SARS- CoV-2 by PCR with specimen collected on or after 3/1/2021

###### Exclusion criteria

- Receipt of a COVID-19 vaccine other than Moderna COVID-19 vaccine on or prior to the date of specimen collection for the SARS-CoV-2 test
- Receipt of 2 doses of Moderna COVID-19 vaccine <24 days apart
- Receipt of a Moderna COVID-19 vaccine <14 days prior to the date of specimen collection for the SARS-CoV-2 test (Day -13 to Day 0, inclusive)
- Receipt more than 2 doses of Moderna COVID-19 vaccine on or prior to the date of specimen collection for the SARS-CoV-2 test
- Having a history of a COVID-19 diagnosis code or SARS-CoV-2 positive molecular test between 12/18/2020 and 2/28/2021, inclusive.

###### Matching

**Test-positive cases:** Individuals who meet the inclusion/exclusion criteria, and have a positive SARS-CoV-2 test on or after 3/1/2021 with the specimen sent for WGS will be defined as cases. For individuals who have more than one positive test after 3/1/2021, sequencing data from the first specimen will be included in analyses. The date of specimen collection for the positive test will be referred to as the index date.

**Test-negative controls:** Individuals who meet the inclusion/exclusion criteria, have a negative SARS-CoV-2 molecular test on or after 3/1/2021, and do not have a positive test on or after 3/1/2021 will be defined as controls. Controls will be randomly selected and matched, in separate processes, for the 2-dose VE analysis and the 1-dose VE analysis.

Individual matching will be applied for both the 2-dose analysis and 1-dose analysis. Controls will be matched to cases by age group (12-17 years [once Moderna COVID-19 vaccine is authorized in this age group], 18-44 years, 45-64 years, 65-74 years, and 75+ years at specimen collection date), sex, race/ethnicity (Non-Hispanic White, Non-Hispanic Black, Hispanic, Non-Hispanic Asian, and Other/Unknown), and specimen collection date (+/- 10 days). The matched date of specimen collection for the negative test will be the index date for the matched control.

###### Outcomes

Variants of concern (VOC) include:

- B.1.1.7 (Alpha)
- B.1.351, B.1.351.2, B.1.351.3 (Beta)
- B.1.617.2, AY.1, AY.2, AY.3, AY.4, AY.5, AY.6, AY.7, AY.8, AY.9, AY.10, AY.11, AY.12

(Delta)

- P.1, P.1.1, P.1.2 (Gamma)

Variants of interest (VOI) include:

- B.1.525 (Eta)
- B.1.526, B.1.526.1, B.1.526.2 (Iota)
- B.1.617.1 (Kappa)
- B.1.617.3

Other variants

- B.1.427, B.1.429 (Epsilon)
- Any other lineage with 15 or more cases
- Lineage unknown

Groupings and naming of VOC and VOI, except Iota, are determined based on SARS-CoV-2 Variant Classifications and Definitions (2021) by Centers for Disease Control and Prevention (CDC) as of 08/24/2021. (Available from: https://[www.cdc.gov/coronavirus/2019-](http://www.cdc.gov/coronavirus/2019-) ncov/variants/variant-info.html). Sub-lineages B.1.526.1 and B.1.526.2 are added to Iota in addition to B.1.526 considering the phylogenetic finding that the B.1.526 lineage is comprised of these two closely related sub-lineages [16] . Updates maybe applied at the time of analysis.

For severe COVID-19 disease, a VOC or VOI with specimen collection date during a hospitalization or with a hospitalization occurring ≤7 days after the specimen collection date (Day 0 to Day 7) will be identified and chart reviewed. COVID-19 hospitalization is defined as hospitalization with severe COVID-19 disease confirmed by chart review. A VOC or VOI with death during the COVID-19 hospitalization is considered a COVID-19 hospitalized death. These outcomes will only be analyzed if samples sizes are sufficient.

###### Exposures

For the 2-dose analysis, the exposure will be defined as receipt of the second dose of Moderna COVID-19 vaccine ≥14 days prior to the SARS-CoV-2 test date vs. not receiving a dose of any COVID-19 vaccine prior to the SARS-CoV-2 test date.

For the 1-dose analysis, the exposure will be defined as receipt of only 1 dose of Moderna COVID-19 vaccine ≥14 days prior to the SARS-CoV-2 test date vs. not receiving a dose of any COVID-19 vaccine prior to the SARS-CoV-2 test date.

###### Statistical Analysis

Separate analyses will be conducted using the TND for 2-dose and 1-dose VE.

For each VOC or VOI, characteristics of test-positive cases and their matched test-negative controls will be described and compared. Specimens with unsuccessful sequencing may also be considered a test-positive case group for analysis. Continuous covariates such as age in years will be summarized by mean, standard deviation, median, quartiles, minimum, and maximum value, and compared using t-test or Wilcoxon rank-sum test, as appropriate; categorical covariates will be summarized by frequency and percentage and compared using chi-square test or Fisher’s exact test, as appropriate. Absolute standardized difference will be calculated to assess the balance of covariates. Potential confounders will be determined based on bivariate analyses and scientific relevance. Given the small sample size and possible need to limit the number of covariates in an adjusted model, potential confounders will be determined by absolute standardized difference (ASD) >0.1 and p-value<0.1, or scientific relevance.

Conditional logistic regression will be used to estimate the odds ratio (OR) and confidence intervals (CIs) of being vaccinated among cases vs controls, without and with adjustment for potential confounders determined from bivariate analyses. VE (%) will be calculated as (1- OR) x 100.

The TND analysis will be performed for both an interim analysis and a final analysis. For each interim analysis and final analysis, the test significance level will be adjusted using the Bonferroni correction, p=0.025 (=0.05/2 tests). As such, we will be calculating 97.5% CIs for each interim and final analysis. We will provide 95% CIs for each analysis as well. If sample

sizes allow, separate analyses using the same methods described above will be considered for VOCs and VOIs associated with severe COVID-19 disease (COVID-19 hospitalization).

For selected VOCs (e.g., Delta variant), if sample sizes allow, separate analyses will be conducted to assess the VE of Moderna COVID-19 vaccine against SARS-CoV-2 variants by time since most recent Moderna COVID-19 vaccination using the methods similar to those described above. Matching possibly will not be maintained in the subgroup analyses. Matching variables (age, sex, and race/ethnicity) and month of specimen collection will be kept in the adjusted model.

#### 10.2 Cohort design

The matched variant cohort will be developed in the same way as the matched cohort assembled for the primary analyses (Section 6.1 and Section 6.2, Study Objectives 1-2), using the same 2-dose Moderna COVID-19 vaccine exposure described in Section 7.1. Because Moderna COVID-19 vaccination began in December 2020 at KPSC, but archiving and WGS of SARS-CoV-2 positive specimens did not begin until March 2021, we will modify the matched cohort from the primary analysis to set index dates to begin no earlier than February 14, 2021 (14 days prior to March 2021). The cohort eligibility criteria will be applied based on the modified index date.

Baseline characteristics of the 2-dose Moderna COVID-19 vaccine cohort will be described and compared between the vaccinated and unvaccinated groups, as for the primary analyses above (Section 8.2). For each VOC or VOI, separately, the incidence of a specific VOC or VOI for the 2-dose vaccinated and unvaccinated individuals will be calculated by dividing the number of individuals with an incident COVID-19 diagnosis with a specific VOC or VOI by the total number of person-years. The person-years for an individual in the cohort will be the time from 14 days after the index date to the date of event of interest, death, disenrollment (allowing for a 31-day gap), end of follow-up (February 28, 2022), or receipt of a dose of any COVID-19 vaccine, whichever comes first. The event date is the date of the first incident COVID-19 diagnosis with a specific VOC or VOI after the index date. In addition, the follow-up time for individuals with an incident COVID-19 diagnosis for which the variant is unknown (i.e., sequencing unsuccessful or not sequenced) or different from the specific variant of interest will be censored on the date of the COVID-19 diagnosis. We will calculate incidence rates of specific VOCs and VOIs, unadjusted and adjusted HRs, and VEs, as described for the primary analyses above (Section 8.3)

If sample size allows, similar analyses will be conducted for VOCs and VOIs associated with COVID-19 hospitalization. Hospitalizations include hospitalized death, but there may be insufficient sample size to assess VE against COVID-19 hospitalized death by variant. Since the cohort analysis will only be conducted at the end of study (February 2022), no adjustment for multiple testing is needed.

#### 10.3 Other considerations

VOCs and VOIs will be selected for analysis based on prevalence in the KPSC population and scientific interest. For a 1:5 matched TND, assuming 60% vaccination rate in test negative

controls, 20 VOC or VOI cases are needed to detect a VE of 0.8 with 80% statistical power. For a 1:1 matched cohort design, approximately 15 VOC or VOI cases are needed in the unvaccinated group to detect a VE of 0.8 with 80% statistical power.

Analyses will be performed at two time points. We will conduct an interim analysis in September 2021 including specimens collected through July 2021 (individuals with VOCs and VOIs and matched test-negative controls). A final analysis will be conducted in April 2022 with specimens collected through February 2022.

### 11 Timelines

##### Table 6. Estimated Project Timelines

| **Description** | **Estimated timeline** |
| --- | --- |
| Contract executed | 01/28/2021 |
| KPSC to submit protocol to Moderna | 02/09/2021 |
| KPSC to obtain IRB approval | 04/30/2021 |
| KPSC to submit final Project Management Plan to Moderna | 05/30/2021 |
| KPSC to submit final Data Management and Statistical Analysis Plan to Moderna | 06/16/2021 |
| Vaccinated individuals accrual | 12/18/2020-12/31/2021 |
| End of follow-up | 12/31/2023 |
| KPSC to submit first interim analysis manuscript to Moderna (Vaccination between 12/18/2020-3/31/2021) | 08/31/2021 |
| KPSC to submit variant VE manuscript to Moderna (interim results) (Specimen collection between March 2021-July 2021) | 10/31/2021 |
| KPSC to submit second interim analysis report (or manuscript if deemed appropriate) to Moderna  (Vaccination between 12/18/2020-6/30/2021) | 11/30/2021 |
| KPSC to submit third interim analysis report (or manuscript if deemed appropriate) to Moderna  (vaccination between 12/18/2020-9/30/2021) | 02/28/2022 |
| KPSC to submit final variant VE manuscript to Moderna (Specimen collection between 03/04/2021-02/28/2022) | 06/30/2022 |
| KPSC to submit fourth interim analysis report (or manuscript if deemed appropriate) to Moderna  (vaccination between 12/18/2020-12/31/2021) | 11/30/2022 |
| KPSC to submit fifth interim analysis report (or manuscript if deemed appropriate) to Moderna  (vaccination between 12/18/2020-12/31/2021) | 05/31/2023 |
| KPSC to submit sixth interim analysis report (or manuscript if deemed appropriate) to Moderna  (vaccination between 12/18/2020-12/31/2021) | 11/30/2023 |
| KPSC to submit end-of-study (final) analysis report tables to Moderna | 09/30/2024 |
| KPSC submission of Final Report to Moderna | 03/31/2025 |
| KPSC to submit final manuscript to Moderna | 6/30/2025 |

### 12 Data Management

Data management activities will be performed by KPSC. The KPSC EHR will be the data source for extracting information on exposures, outcomes, and covariates. KPSC will develop the study datasets, maintain documentation, and perform data quality checks.

Programmers will extract data per protocol and reference documentation for KPSC EHR databases. Study cohorts and outcomes will be double programmed by two programmers independently. Programmers will conduct data quality checks which include data integrity control and program review. Data integrity control will include checks such as sample size, duplications, formatting, etc. All decisions made during data extraction and data quality checks will be documented.

### 13 Limitations

This study has several potential limitations. Misclassification of the exposure is possible if COVID-19 vaccines were received outside of the KPSC system. However, external COVID-19 vaccination can be captured in the KPSC EHR data by manual entry and electronic updates from the California Immunization Registry (CAIR) (for which KPSC has a proactive mechanism in place for obtaining regular updates from CAIR). Misclassification of COVID-19 outcomes is also possible due to imperfect capture and sensitivity of SARS-CoV-2 molecular diagnostic tests. For variant VE analyses (Secondary Objectives 16-17), sequencing may be unsuccessful if viral loads are low, which may be more common among vaccinated vs. unvaccinated individuals. However, samples with unsuccessful sequencing may also be considered a test- positive case group for analysis. For patients with COVID-19 symptoms, sensitivity of molecular diagnostic tests is generally high (>90%). Efforts are made to ask patients about positive tests conducted outside of KPSC and to document in the EHR with internal diagnosis codes. In addition, we will be unable to capture some potential confounders, such as occupational risk exposures, behavioral factors (e.g., masking, distancing, handwashing), and residential factors (e.g., congregate settings) that impact an individual’s risk for COVID-19. Thus, despite all efforts to ensure comparability, there may be some unmeasured differences between vaccinated and unvaccinated cohorts. Some secondary analyses may be underpowered; their meaningfulness will depend on uptake of Moderna COVID-19 vaccine in these patient populations.

### 14 Human Subjects Protection

The study will be reviewed and approved by the KPSC Institutional Review Board (IRB). All study staff with access to protected health information are trained in procedures to protect the confidentiality of subject data. We will obtain a waiver of informed consent as this is an observational study of authorized and recommended Moderna COVID-19 vaccine administered in the course of routine clinical care.

The HIPAA Privacy Rule governs the use and disclosure of personally identifiable information (protected health information; PHI) from covered entities. Throughout the course of this study, no PHI will be disclosed, however it will be accessed through the EHR. This information will only be accessed by those authorized to do so and will not be shared with Moderna or anyone outside of the KPSC study team. As this access presents no more than minimal risk to individuals and the research could not be practically done if required to obtain written authorization for usage, we will obtain a waiver for written HIPAA authorization for research involving use of the EHR.

*B.1.526 Identified in New York.* medRxiv, 2021.
